# Large-scale brainstem neuroimaging and genetic analyses provide new insights into the neuronal mechanisms of hypertension

**DOI:** 10.1101/2023.10.25.23297515

**Authors:** Tiril P. Gurholt, Torbjørn Elvsåshagen, Shahram Bahrami, Zillur Rahman, Dennis van der Meer, Alexey Shadrin, Oleksandr Frei, Tobias Kaufmann, Ida E Sønderby, Sigrun Halvorsen, Lars T. Westlye, Ole A. Andreassen

## Abstract

**Background:** While brainstem regions are central regulators of blood pressure, the neuronal mechanisms underlying their role in hypertension remain poorly understood. Here, we investigated the structural and genetic relationships between global and regional brainstem volumes and blood pressure.

**Methods:** Using magnetic resonance imaging data from n=32,666 UK Biobank participants, we assessed the association of volumes of the whole brainstem and its main regions with blood pressure. We applied powerful statistical genetic tools, including bivariate causal mixture modeling (MiXeR) and conjunctional false discovery rate (conjFDR), to summary statistics from genome-wide association studies of brainstem volumes (n=27,034) and blood pressure (n=318,891) and evaluated their genetic architectures.

**Results:** We observed negative associations between the whole brainstem and medulla oblongata volumes and systolic blood and pulse pressure, and positive relationships between midbrain and pons volumes and blood pressure traits when adjusting for the whole brainstem volume (all ∣r∣ 0.03-0.05, p≤0.0042). We observed the largest genetic overlap for the whole brainstem, sharing between 54% and 69% of its trait-influencing variants with blood pressure. We identified 42 shared loci between brainstem volumes and blood pressure traits and mapped these to 83 genes, implicating molecular pathways linked to the development of the brainstem, cranial nerves, and sympathetic neurons, and to metal ion transport and cell-matrix adhesions.

**Conclusions:** The present findings support a link between brainstem structures and blood pressure and provide new insights into their shared genetic underpinnings. The overlapping genetic architectures and mapped genes offer new mechanistic information about the roles of brainstem regions in hypertension.

## Introduction

Hypertension is one of the leading contributors to ischemic heart disease, heart failure, and stroke and accounts for at least 13% of deaths worldwide.^1–4^ The pathophysiology of chronic blood pressure elevation remains poorly understood, and only half of those with hypertension have adequate blood pressure control.^5,6^

The brainstem is a regulator of blood pressure^7,8^ and includes the midbrain, pons, and medulla oblongata. Disentangling the role of the brainstem in the neuronal networks underlying human blood pressure regulation is one critical step toward more effective treatments and improved outcomes in hypertension. The evidence for neuronal involvement in blood pressure regulation comes primarily from animal models.^9^ Invasive animal studies suggest that nuclei within brainstem regions have central roles in regulating sympathetic nerve activity, which is a major contributor to the hypertensive state.^9–11^ Of particular interest for understanding sympathetic nerve tone are neurons within the rostral ventrolateral medulla oblongata and midbrain; these are critical for experimental hypertension in rodents.^8,12^ We know less about the roles of brainstem regions in human blood pressure regulation. Some recent functional magnetic resonance imaging (MRI) studies assessed brainstem nuclei activity in humans in response to pharmacological stimulation, orthostatic stress, and hypoxia and suggested that several nuclei are part of the central cardiovascular network.^13–15^ Yet, there is no previous investigation of whether and how the structure of these regions influences blood pressure in humans.

The unprecedented availability of a large neuroimaging dataset from the UK Biobank^16^ and the development of a brainstem segmentation algorithm^17^ allowed us to identify the first genetic loci linked to the structure of individual brainstem regions through genome-wide association studies (GWAS).^18^ Prior GWASs have also shed light on the genetic underpinnings of human blood pressure,^19–21^ with the most recent study identifying 505 independent loci linked to blood pressure traits.^21^ Despite the substantial genetic contributions to brainstem structure and blood pressure variation and their putative mechanistic links, the genetic relations between brainstem regions and blood pressure traits are largely unknown.

The present study investigated the phenotypic and genetic relationships between global and sub-regional brainstem volumes and blood pressure. To this end, we tested the associations between MRI-based brainstem volumes and blood pressures of UK Biobank participants (n=32,666). We performed follow-up analyses for age- and sex-related patterns. Next, to characterize the overlap in genetic architectures we applied the bivariate causal mixture model (MiXeR)^22,23^ and the conjunctional false discovery rate (conjFDR) method^24,25^ to summary statistics of previous GWASs of brainstem volumes (n=27,034)^18^ and blood pressure (n=318,891).^21^ MiXeR estimates the total number of trait-influencing variants for a given trait and the total number of shared and unique trait-influencing variants for a pair of traits.^22,23^ ConjFDR enables the detection of individual genetic loci jointly associated with two phenotypes.^24,25^

## Methods

### Study design and participants

For the structural analyses, we included 32,666 UK Biobank participants (53.8% women) with brain MRI and blood pressure data. We excluded participants who did not pass quality control (see below) or withdrew their informed consent (opt-out list dated August 9^th^, 2021). The UK Biobank obtained informed consent from all participants, has IRB approval from the North West Multi-center Research Ethics Committee, and the Ethics Advisory Committee (https://www.ukbiobank.ac.uk/ethics) oversees the UK Biobank Ethics & Governance Framework.^16^ We obtained access to the UK Biobank cohort through Application number 27412. We have approval from the Regional Committees for Medical and Health Research Ethics in Norway.

For the genetic analyses, we included GWAS summary statistics of blood pressure traits of 318,891 participants from the Trans-ancestry Million Veteran Program (MVP) consortium accessed through dbGAP under accession number phs001672.v9.p1.^21^ The MVP consortium obtained consent from all participants and followed all relevant ethical regulations.^21^ We included our previous GWAS summary statistics of brainstem volumes of 27,034 white European UK Biobank participants.^18^

### Demographic and clinical data

We extracted demographic data (age, sex, self-reported ethnicity) and systolic and diastolic blood pressure from the UK Biobank repository. We extracted potential confounders and relevant phenotypic variables, e.g., based on our previous research,^26^ including body anthropometrics (waist circumference, hip circumference, body mass index (BMI)), self-identified ethnic ancestry, and variables reflecting cardiovascular risk (i.e., self-reported history of diabetes, hypercholesterolemia, hypertension, current cigarette smoker, and current alcohol consumption).

For blood pressure, we computed the average of two subsequent automatic blood pressure readings when available for systolic and diastolic blood pressure. If only a single automatic blood pressure reading were available, we used the single measurement. If no automatic readings were available, we would similarly use the manual blood pressure assessment. Subsequently, we computed pulse pressure as the difference between systolic and diastolic blood pressure. Additionally, we computed waist-to-hip ratio (WHR) as waist circumference divided by hip circumference. See **Note S1** for the extracted UK Biobank data-field IDs and additional data processing details.

### MRI acquisition, processing, and quality control

The participants’ inclusion was performed on three different sites (Cheadle, Reading, or Newcastle), and all participants underwent 3T brain MRI using similar scanners and protocols.^16^ All data for each participant, including MRI and blood pressure traits, were collected on the same day and site.

We have previously described the processing of brain MRI in detail.^18^ Briefly, we processed brain MRI DICOM data *in-house* using FreeSurfer^27^ (version 5.3.0; http://www.freesurfer.net), followed by Bayesian brainstem segmentation and delineation of the whole brainstem, midbrain, pons, superior cerebellar peduncle, and medulla oblongata using FreeSurfer (version 6.0).^17^ We extracted the whole brainstem, midbrain, pons, and medulla oblongata and estimated intracranial volume (ICV),^28^ and additionally, Euler numbers^29^ as a proxy for image quality,^30^ for the statistical analyses.

We applied MRI quality assessment using Euler numbers.^30^ Among the 42,063 participants with available brain MRI, we removed three with missing Euler numbers and 4,206 based on a previously outlined procedure.^26^ Second, we removed the remaining participants who did not pass manual MRI quality control,^18^ further excluding 48 participants. Lastly, we removed 5,140 participants with incomplete demographic and clinical data needed for the statistical analyses. In turn, this resulted in the final sample of 32,666 participants.

### Statistical analysis of brainstem volumes

We investigated the sample demographics and brainstem volumes across and within sexes. We compared categorical variables using the χ^2^-test. For normally or non-normally distributed continuous variables, we used the two-sample t-test or the Wilcoxon rank-sum test, respectively. For unequal variance across sexes, we replaced the t-test with the Welch approximation. We verified normality by visually evaluating density plots (**Figures S1-S2**).

We used multiple linear regression to investigate the association between blood pressure traits and the whole brainstem and its main regions. We adjusted for age, age^2^, sex, age-by-sex, age^2^-by-sex, WHR, WHR^2^, ethnicity, and metabolic/lifestyle variables (including current cigarette smoking (yes/no), current alcohol consumption (yes/no), diabetes (yes/no), and hypercholesterolemia (yes/no)), ICV, image quality (average Euler number), and site. We adjusted for Age^2^ to account for age-related nonlinearities, and WHR and WHR^2^ since we have previously shown that WHR is nonlinearly associated with the whole brainstem.^26^ The analyses for brainstem regions were additionally adjusted for whole brainstem volume, thus revealing associations with blood pressure beyond global brainstem volume, analogous to recent studies.^18,31,32^ We entered categorical variables as factors and mean-centered all continuous variables.

We performed follow-up analyses in the participants with a normal blood pressure range, i.e., excluding those with hypertension (here defined as self-reported hypertension or systolic blood pressure ≥140 mmHg or diastolic blood pressure ≥90 mmHg on the day of study participation). Lastly, we investigated age- and sex-related patterns by conducting separate analyses for participants aged <60 years and for participants aged ≥60 years and for men and women. The supplement includes complementary analyses of brainstem regions without adjustment for whole brainstem volume (**Note S2**).

We performed statistical analyses in R (version 3.6.0; https://www.r-project.org). We computed the partial correlation coefficients, *r*, effect size directly from the t-statistics for continuous variables and via Cohen’s d for categorical variables.^33^ We evaluated model residuals for normality using residual vs. fitted value and quantile-quantile (Q-Q) plots and decided to move forward without log-transformation of any regression model variables. We used Bonferroni correction to adjust for multiple comparisons at α=0.05 across N_1_=12 independent tests; i.e., the number of linear regression models, counting partly overlapping models once, reflecting the included four brain structures and three blood pressure traits, yielding a significance threshold of *p*≤α/N_1_=0.0042 (rounded to four decimal points) for the brainstem volume analyses.

### Genetic overlap analyses and functional annotation

To perform genetic overlap and functional annotation analyses, we used the available GWASs summary statistics of blood pressure traits (n=318,891)^21^ and brainstem volumes (n=27,034).^18^

First, we applied the linkage disequilibrium (LD) score regression (LD Score v1.0.1) method^34,35^ to determine the genetic correlation (r_g_) between summary statistics of blood pressure traits and brainstem volumes. Subsequently, we applied the bivariate causal mixer model (MiXeR)^22,23^ to the same GWAS summary statistics. Briefly, MiXeR estimates the total number of shared and trait-specific trait-influencing SNPs and SNP-based heritability (h^2^_snp_) for each trait based on the distribution of z-scores and detailed modeling of the LD structure. We present the results as Venn diagrams, illustrating the proportion of shared and trait-specific trait-influencing SNPs. We used the Akaike Information Criterion (AIC), where a positive AIC indicates adequate discrimination between the fitted and the comparative model.^23^ We evaluated the model fit based on predicted versus observed conditional Q-Q plots (a closer fit between the dashed and continuous line of the same color suggests a better fit) and log-likelihood plots that visualize the parameter estimation procedure (**Figures S3-S5**).

Following MiXeR, we used conjFDR^24,25^ to determine shared genetic loci between traits. Briefly, conjFDR is the maximum of two conditional FDR (condFDR) statistics of a given SNP, where one trait is conditioned on the other and vice versa. It yields an estimate of the posterior probability that a SNP is null for either or both traits, provided that the p-values for both phenotypes combined^25^ are equal to or smaller than the p-values for each individual trait. We considered loci significant if the lead SNP passed the genome-wide significance threshold. To avoid potential biases due to complex LD structures, we performed the analysis after excluding SNPs in the extended major histocompatibility complex (MHC; hg19 location chr 6: 25119106-33854733) and 8p23.1 (hg19 location chr 8: 7242715-12483982) regions.

Lastly, we used the Open Targets platform to map the significant SNPs to genes. Open Targets maps SNPs to genes by combining positional information regarding the distance between the variant and each gene’s canonical transcription start site, eQTL, pQTL, splicingQTL, epigenomic data, and functional prediction.^36^ After excluding genes in the MHC region, we applied gene-set analyses with a Bonferroni correction on all mapped genes using FUMA.^37^

## Results

### Demographic and clinical data

**Table S1** summarizes the demographic and clinical data. The final sample (n=32,666) included more women (n=17,561; 53.8%) than men (n=15,105; 46.2%) and was mainly of self-identified white European ancestry (97.0%). The age range was 44-82 years; on average, the women were slightly younger than men (mean±std ages 62.5±7.3 and 63.6±7.6 years, respectively). Men had, as expected,^26^ generally higher levels of adverse cardiometabolic factors than women.

### Brainstem volumes and blood pressure traits

Men had larger volumes of the whole brainstem and brainstem regions and higher systolic and diastolic blood pressure and pulse pressure than women (**Table S2**; **Figure S2**). The analysis revealed significant negative associations between the whole brainstem volume and pulse pressure (r=-0.03, p=5.1×10^−9^) and systolic blood pressure (r=-0.02, p=6.4×10^−5^) (**Figure 1a**; **Table S3a**). Medulla oblongata volume was negatively associated with pulse pressure (r=-0.02, p=7.8×10^−6^) and diastolic (r=-0.03, p=1.7×10^−7^) and systolic (r=-0.04, p=2.4×10^−10^) blood pressure. Pons volume was positively associated with pulse pressure (r=0.02, p=0.0004) and diastolic (r=0.02, p=0.0040) and systolic (r=0.02, p=2.1×10^−5^) blood pressure. Midbrain volume was positively associated with systolic (r=0.02, p=0.0010) and diastolic (r=0.02, p=0.0018) blood pressure (**Figure 1b**; **Table S3b**). We observed no other significant associations between brainstem volumes and blood pressure traits.

**Figure 1:**
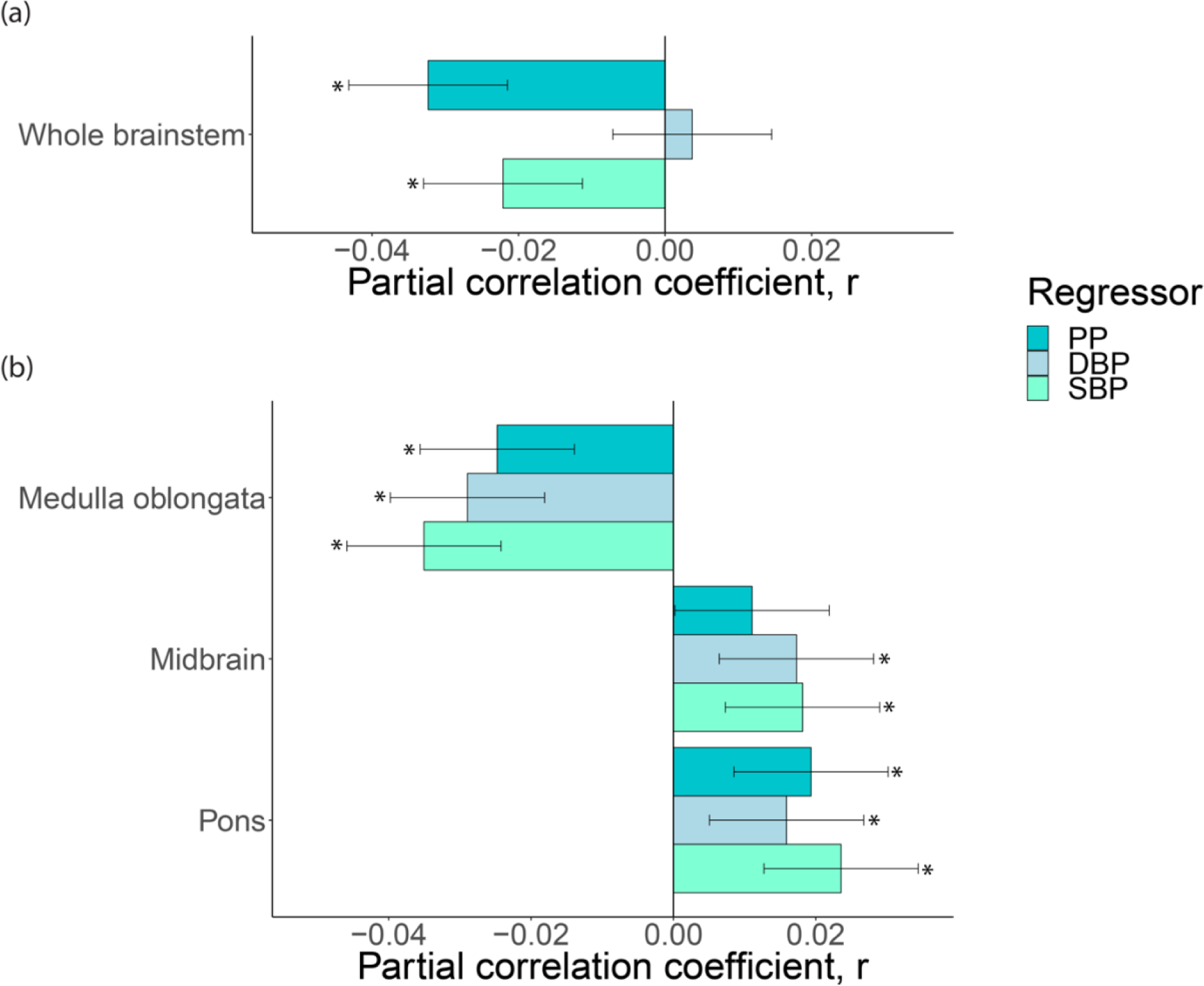
Blood pressure and brainstem volumes. *Notes:* The association between blood pressure metrics and volumes of the **(a)** whole brainstem and **(b)** brainstem regions (n=32,666). We adjusted for covariates as outlined in the Methods section. *Abbreviations: PP – pulse pressure; DBP – diastolic blood pressure; SBP – systolic blood pressure*.

In follow-up analyses of non-hypertensive participants (n=15,870; 61.4% women), we found a similar association pattern as in the full sample, albeit with fewer significant findings, likely due to the smaller sample (**Figure S6**; **Table S4ab**). Moreover, when the analyses for brainstem regions were run without adjusting for the whole brainstem, there were significant negative associations between medulla oblongata volumes and systolic blood pressure (r=-0.04, p=1.4×10^−12^) and pulse pressure (r=-0.04, p=1.4×10^−13^), pons volume was negatively associated with pulse pressure (r=-0.03: p=1.0×10^−6^) and systolic blood pressure (r=-0.02, p=0.0035), and midbrain volume was negatively associated with pulse pressure (r=-0.02, p=2.6×10^−5^).

### Age-related patterns

For those aged 60 and above (n=21,697; 52.2% women), we observed similar significant relationships between brainstem volumes and blood pressure traits as in the primary analyses (**Figure S7ab**; **Table S5ab).** In those under 60 (n=10,969; 57.0% women), we observed a similar pattern of relationships but no significant findings (**Figure S7cd**; **Table S6ab**).

### Sex-related patterns

We observed similar relationships between the whole brainstem volume and blood pressure traits in women (n=17,561) and men (n=15,105), with significantly lower volume at higher pulse pressure (r=-0.04 and p=1.8e-07 for women and r=-0.03 and p=0.0008 for men) (**Figure 2**; **Tables S7ab-8ab**). For brainstem regions, women showed significantly lower medulla oblongata volume at higher pulse pressure (r=-0.05, p=2.3e-09), diastolic (r=-0.03, p=0.0006), and systolic (r=-0.05, p=1.9e-10) blood pressure, higher midbrain volume at higher systolic blood pressure (r=0.02, p=0.0020) and pulse pressure (r=0.02, p=0.0026, and higher pons volume at higher pulse pressure (r=0.03, p=5.8e-05) and systolic blood pressure (r=0.03, p=2.6e-05). For men, our findings suggest significantly lower medulla oblongata volume at higher diastolic blood pressure (r=-0.03, p=0.0001) and higher midbrain volume at higher diastolic blood pressure (r=0.02, p=0.0034).

### Genetic overlap analyses and functional annotation

First, we found a significant positive genetic correlation between the whole brainstem and diastolic blood pressure (r_g_=0.109, SE=0.048, p=0.024; **Figure S8**) using LD Score regression but observed no other significant genetic correlations.

Second, using univariate MiXeR, we observed that blood pressure is more polygenic than the brainstem. As illustrated by the Venn diagrams (**Figures 3-5**), the number of trait-influencing variants for systolic blood pressure, diastolic blood pressure, and pulse pressure were up 4-6 times as high (were 3.2K, 2.8K, and 2.1K, respectively) as for brainstem regions (0.5K-1.3K).

**Figure 3:**
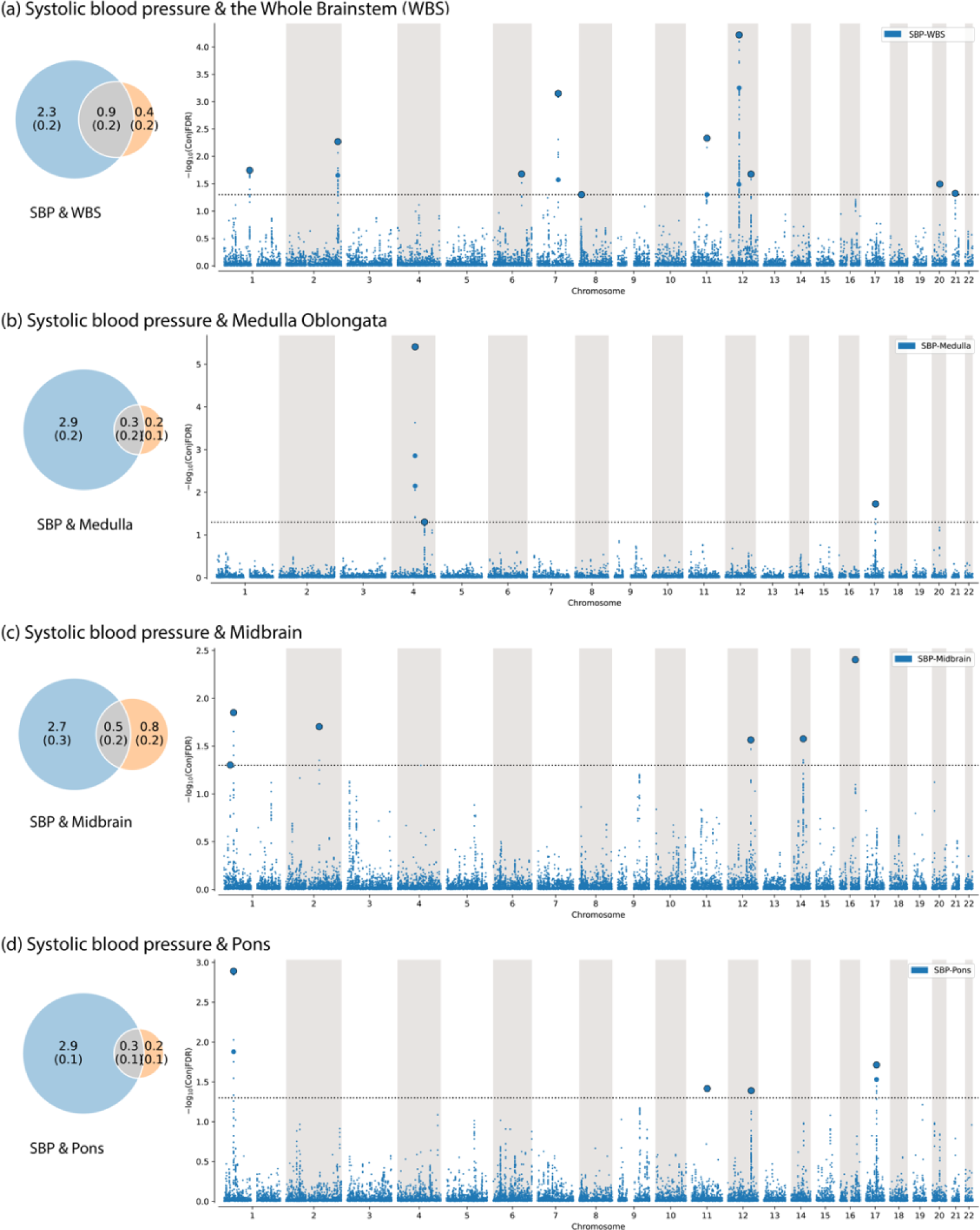
Genetic overlap between systolic blood pressure and brainstem volumes. *Notes:* The figure shows the (***left***) Venn diagrams of the polygenicity of blood pressure metrics (blue) and brainstem regions (orange) and the polygenic overlap between traits (gray); and (***right*** Manhattan plots of shared loci - between pulse pressure and (a) the whole brainstem, (b) Medulla oblongata, (c) Midbrain, and (d) Pons. *Abbreviations: Medulla – Medulla Oblongata; SBP – systolic blood pressure; WBS – whole brainstem*.

The bivariate MiXeR analysis revealed polygenic overlap between brainstem regions and blood pressure. The highest estimated number of shared trait-influencing variants are between the whole brainstem and systolic blood pressure (0.9K; i.e., 69% of the entire brainstem variants), diastolic blood pressure (0.8K; i.e., 61% of the whole brainstem variants), and pulse pressure (0.7K; i.e., 54% of the whole brainstem variants), as illustrated by the intersection of corresponding Venn diagrams (**Figures 3-5**). We observed the second highest estimated number of shared trait-influencing variants between the midbrain and systolic blood pressure (0.5K; i.e., 38% of the midbrain variants), diastolic blood pressure (0.5K; i.e., 38% of the midbrain variants), and pulse pressure (0.4K; i.e., 31% of the midbrain variants). The estimated numbers for trait-influencing variants shared between the pons and medulla oblongata and blood pressure traits were lower and ranged between 0.1K and 0.3K.

Third, we observed successive increments of SNP enrichment for the whole brainstem and its regions when conditioned on association p-values for blood pressure traits on conditional Q-Q plots of the condFDR analysis (**Figures S9-S11**). The consistently increasing leftward deflection for subsets of variants with higher significance in the conditional trait in both directions indicates substantial polygenic overlap between the brainstem and blood pressure traits.

Fourth, we revealed 42 loci shared between the brainstem volumes and blood pressure traits using the conjFDR analysis (**Tables S9-S33**) and illustrated the observed genetic overlaps using Manhattan plots (**Figures 3-5**). We identified 18, 14, and 31 shared loci between systolic blood, diastolic blood, and pulse pressure, and the whole brainstem and its regions, respectively. Specifically, we identified (i) ten, six, four, and three loci shared between systolic blood pressure and the whole brainstem, midbrain, pons, and medulla oblongata, respectively (**Figure 3**); (ii) seven, five, one, and two loci were jointly associated with diastolic blood pressure and the whole brainstem, midbrain, pons, and medulla oblongata, respectively (**Figure 4**); and (iii) 22, seven, eight, and three loci were shared between pulse pressure and the whole brainstem, midbrain, pons, and medulla oblongata, respectively (**Figure 5**). For both systolic and diastolic blood pressure, we observed the overall most significant genetic overlaps for medulla oblongata on chromosome 4, followed by the whole brainstem and chromosome 12. For pulse pressure, we observed the overall most significant results for pons followed by medulla oblongata on chromosome 17, and the overall largest number of shared loci for the whole brainstem, with the most significant findings for chromosomes 7 and 20.

**Figure 4:**
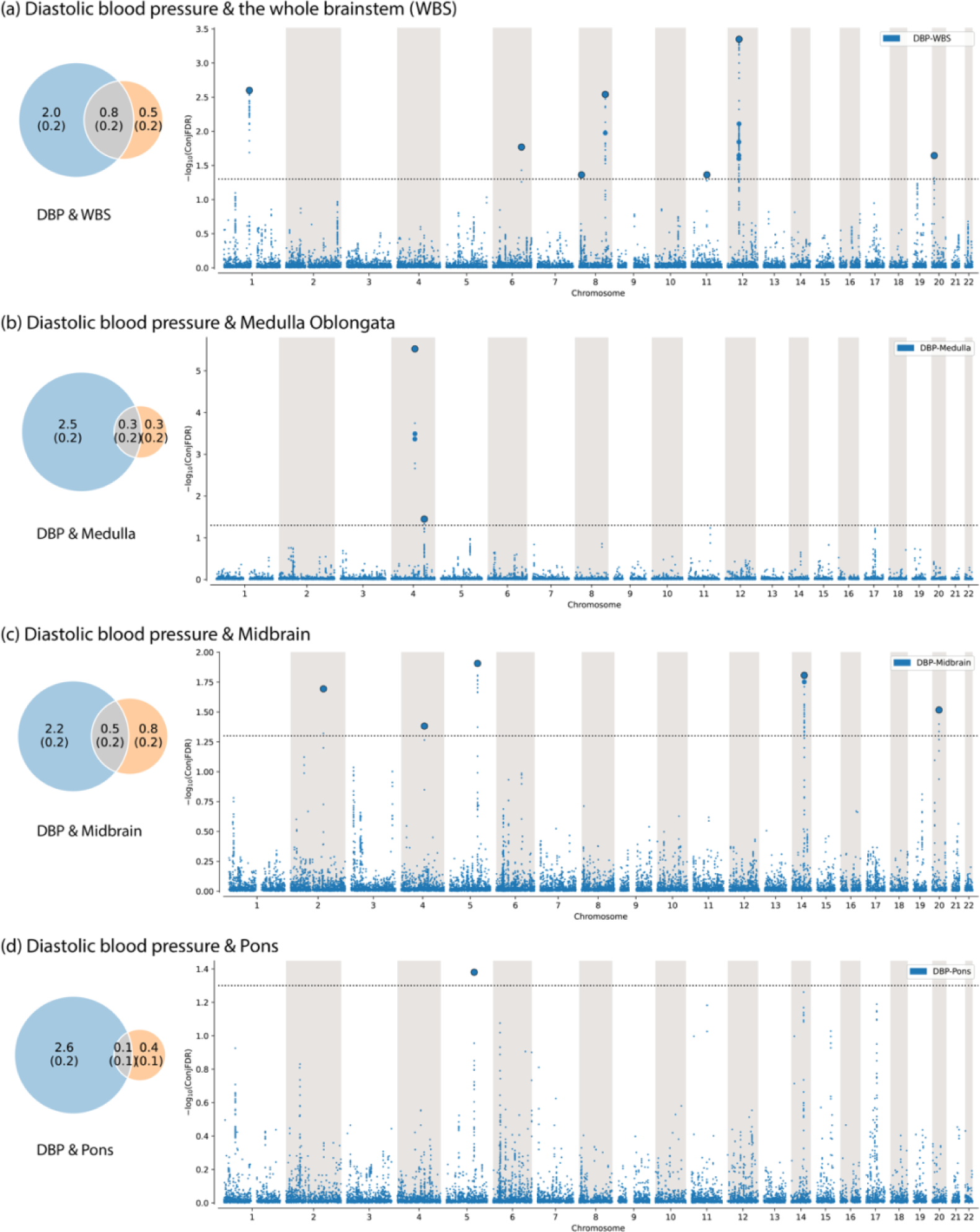
Genetic overlap between diastolic blood pressure and brainstem volumes. *Notes:* The figure shows the (***left***) Venn diagrams of the polygenicity of blood pressure metrics (blue) and brainstem regions (orange) and the polygenic overlap between traits (gray); and (***right*** Manhattan plots of shared loci - between pulse pressure and (a) the whole brainstem, (b) Medulla oblongata, (c) Midbrain, and (d) Pons. *Abbreviations: DBP – diastolic blood pressure; Medulla – Medulla Oblongata; WBS – whole brainstem*.

**Figure 5:**
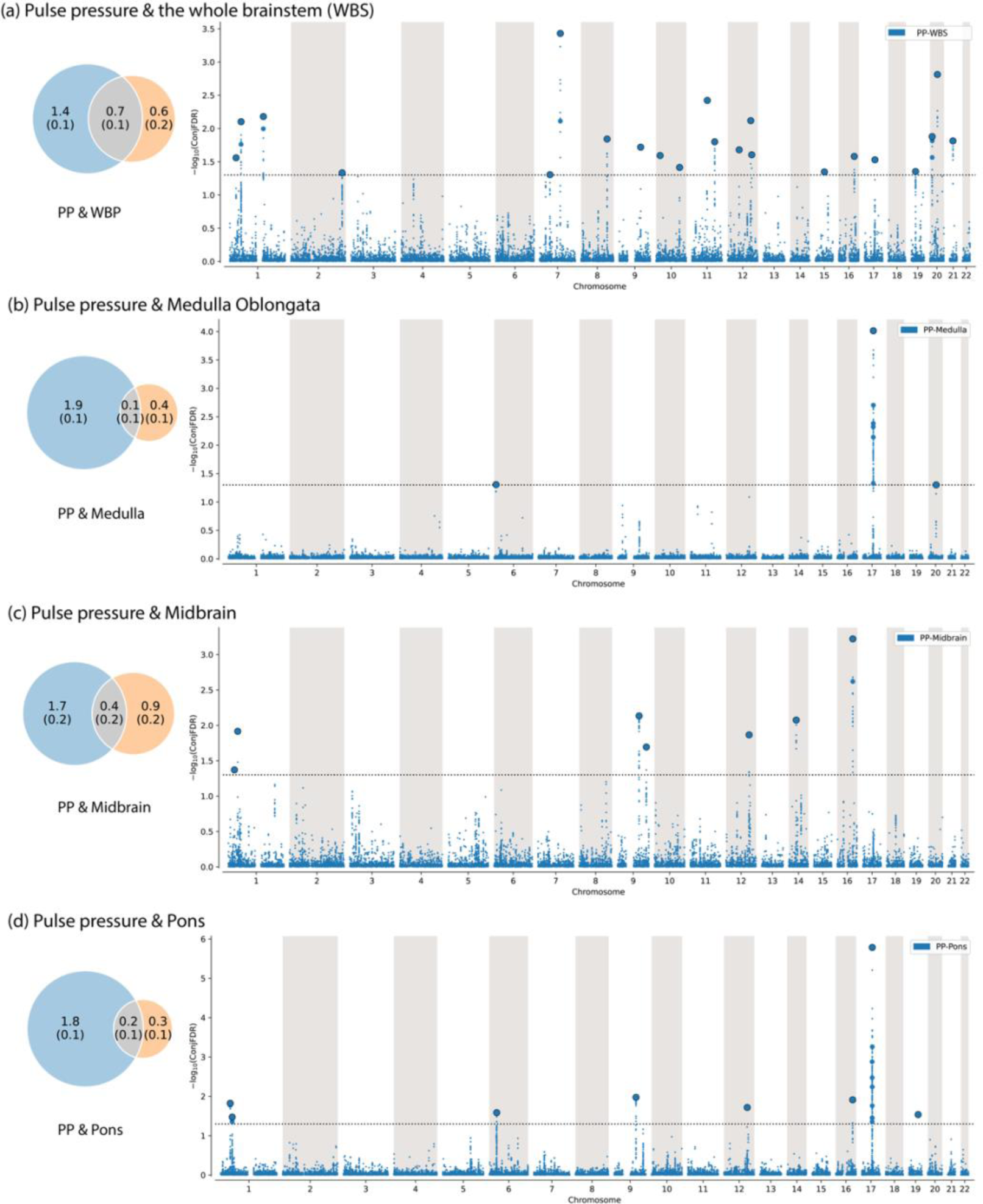
Genetic overlap between pulse pressure and brainstem volumes. *Notes:* The figure shows the (***left***) Venn diagrams of the polygenicity of blood pressure metrics (blue) and brainstem regions (orange) and the polygenic overlap between traits (gray); and (***right*** Manhattan plots of shared loci - between pulse pressure and (a) the whole brainstem, (b) Medulla oblongata, (c) Midbrain, and (d) Pons. *Abbreviations: Medulla – Medulla Oblongata; PP – pulse pressure; WBS – whole brainstem*.

Lastly, we mapped all lead SNPs for each shared locus from conjFDR analyses to genes using the Open Targets platform (**Tables S9-S33**). We observed 33, 24, and 57 mapped genes between systolic blood, diastolic blood, and pulse pressure with brainstem regions, respectively (**Tables S34-S35**). Subsequently, we performed gene-set analyses on all the mapped genes. We observed significant enrichment for 3, 27, and 12 gene set terms when mapped with genes for pulse pressure and the whole brainstem, medulla oblongata, and pons, respectively (**Tables S36-S38**). The enriched gene-sets for pulse pressure and the whole brainstem reflect processes related to cell differentiation and cell-matrix adhesions. In contrast, the enriched gene-sets for pulse pressure and medulla oblongata and pons identified highly expressed *HOXB1, HOXB2, HOXB3, HOXB4,* and *BMP6* genes, and reflect different biological processes such as the development of rhombomeres and cranial nerves (**Tables S36-S38**). There was no significantly enriched gene-set for systolic and diastolic blood pressure and brainstem volumes.

## Discussion

While the brainstem plays a known role in hypertension, how its structure and genetic architecture are linked to human blood pressure remains largely unknown. Here we identified negative associations between the whole brainstem and medulla oblongata volumes and blood pressure traits and positive relationships between midbrain and pons volumes and blood pressure when adjusting for whole brainstem volume, leveraging the UK Biobank participants and powerful analytical tools. We also discovered extensive genetic overlap for whole brainstem volume and blood pressure traits, which shared 54% - 69% of the brainstem’s trait-influencing variants. Finally, we identified 42 individual genetic loci jointly associated with brainstem structures and blood pressure traits and mapped these to 83 genes implicating molecular pathways linked to brainstem and cranial nerve development, cell differentiation, and cell-matrix adhesions.

The current study is the first large-scale neuroimaging study linking blood pressure to brainstem structure variation. We observed a consistent pattern linking brainstem volume to blood pressure traits both in middle age and in late-life. Although some previous work reported positive associations between blood pressure traits and late-life brain volumes,^38,39^ most studies observed negative associations between brain volumes and blood pressure across adulthood.^40–43^ These found that higher blood pressure and hypertension were related to lower total brain volume and reduced regional brain volumes, including smaller frontal and medial temporal structures, e.g., the hippocampus.^40–43^ Furthermore, some studies found that higher blood pressure in early adulthood and midlife was associated with smaller late-life brain volumes.^44,45^ Our findings suggest a continuous association with small effects between brainstem volumes and blood pressure in non-hypertensive and hypertensive states.

Furthermore, we observed indications of different association patterns between brainstem regions and blood pressure traits in women and men, with predominantly significant associations in women but not men, despite, on average lower blood pressure, general cardiometabolic risk, and age for women. Indeed, our observations may relate to sex differences in blood pressure regulation, hypertension, and cardiovascular disease, as well as sex hormones^46–48^ and, e.g., body composition, lifestyle, and environmental factors.^48^ To address these observed sex differences fully and how they relate to hypertension disease risk and related markers and outcomes, designated studies are required.

MiXeR revealed substantial genetic overlap between brainstem volumes and blood pressure. We found the largest number of shared trait-influencing variants for the whole brainstem, which shared between 54% and 69% of its trait-influencing variants with blood pressure traits. For the brainstem regions, the midbrain showed the greatest overlap, which shared 31% to 38% of its trait-influencing variants with blood pressure traits. Despite the substantial genetic overlap, the genetic correlations between brainstem volumes and blood pressure traits were weak (all ∣r∣ < 0.11). This pattern of genetic overlap with weak correlation suggests mixed allelic effect directions since the consistent direction of effect across overlapping genetic loci is required for a significant genetic correlation.^49^ In support of this notion, we found that 50% of the 42 individual genetic loci jointly associated with brainstem volumes and blood pressure traits had concordant effects (e.g., greater volume and higher blood pressure). The remaining 50% of the shared loci exhibited discordant effects (e.g., greater volume and lower blood pressure).

The significant associations between the brainstem volumes and blood pressure metrics in the UK Biobank participants and the substantial genetic overlap revealed by MiXeR provide further support for the relevance of brainstem regions in blood pressure regulation. Although the precise neural networks and roles of brain regions in cardiovascular regulation remain to be fully clarified, a large body of evidence implicates several nuclei within the medulla oblongata, midbrain, and pons in blood pressure regulation.^7,9,12^ The rostral ventrolateral medulla oblongata is a key regulator of blood pressure and includes the glutamatergic C1 neurons.^7^ These neurons produce adrenaline and project to preganglionic sympathetic and parasympathetic neurons, which in turn innervate the heart, arterioles, and kidneys.^7–9^ The C1 neurons are thus likely contributors to the excessive sympathetic nerve activity commonly found in hypertension.^7,9,12^ Rostral ventrolateral medulla oblongata neurons also project to and receive inputs from other regions within the brainstem linked to blood pressure regulation, such as the periaqueductal gray matter of the midbrain and the locus coeruleus of the pons.^9^ An involvement of midbrain and pontine nuclei in blood pressure regulation is also supported by invasive electrical stimulation studies in individuals with movement disorders; here, electrical stimulation of midbrain^50^ and pontine^51^ nuclei modulated blood pressure.

The current findings implicate novel molecular pathways in hypertension and several of the shared loci identified by the conjFDR method and the mapped genes are noteworthy. The most significant shared locus for systolic and diastolic blood pressure - rs13107325 - was jointly associated with medulla oblongata volume. rs13107325 is in the *SLC39A8* gene and encodes a cellular metal ion transporter. The major allele of *SLC39A8* is highly pleiotropic, associated with increased systolic and diastolic blood pressure,^52^ and is essential for normal heart development.^53^ However, the precise mechanisms by which the *SLC39A8* influences blood pressure remain to be clarified. rs76169231 and rs11111293 were jointly associated with systolic blood pressure and pulse pressure and the whole brainstem, midbrain, and pons. These two loci were mapped to the *WASHC3* gene. *WASHC3* regulates endolysosomal membrane trafficking^54^ and how the gene may contribute to hypertension mandates further research. We also found that several of the loci jointly associated with brainstem volumes and blood pressure were mapped to *HOX* genes. Furthermore, *HOX* genes were included in most of the significantly enriched gene sets for pulse pressure and brainstem volumes. *HOX* genes encode transcription factors with central roles in nervous system development,^55^ and the *HOXB1-4* genes are critical for the development of the pons and the medulla oblongata.^56^ Interestingly, *HOXB* genes are involved in developing sympathetic neurons, yet how these genes relate to blood pressure regulation remains elusive. The *BMP6* gene was also represented in several of the significantly enriched gene sets. The *BMP* genes are involved in sympathetic nervous system development^57^ and are increasingly recognized for their roles in cardiovascular homeostasis.^58^ The gene-set analyses for pulse pressure and brainstem volumes also identified molecular pathways linked to cranial nerve development, cell differentiation, and cell-matrix adhesions. Further experimental studies of the overlapping genetic pathways are needed to determine their precise molecular contributions to hypertension.

The MRI segmentation algorithm used in the present study delineates larger regions of the brainstem that each include a substantial number of nuclei – of which only a subset has been linked to blood pressure regulation – and white matter.^17^ Indeed, this is one potential explanation for why the observed associations between brainstem volumes and blood pressure in the UK Biobank participants were small, with significant effect sizes ranging from ∣r∣ 0.03 to 0.05. However, new methods for analyzing individual brainstem nuclei were recently developed,^59,60^ and stronger associations might be revealed in future studies of brainstem structure and blood pressure.

The present study has several strengths and limitations. Concerning strengths, the current work includes a large sample of genotyped and well-characterized participants with brain MRI and blood pressure data from the UK Biobank. This allowed us to adjust the analyses for several potentially confounding factors, such as body anthropometrics and other cardiovascular risk factors. Furthermore, we used summary statistics from the largest GWASs of blood pressure and brainstem volumes published to date and employed a recently developed and robust method for segmenting brainstem regions. When it comes to the limitations, the UK Biobank sample is of largely self-identified white European origin and does not reflect the ethnic diversity of the general population. Moreover, the GWAS of blood pressure traits included a sample with mixed ancestry (69.1% non-Hispanic whites) and predominantly male (91.5%) participants.^21^ In comparison, the GWAS of brainstem volumes was restricted to a sample of individuals with white European ancestry and a balanced sex distribution (52% women).^18^ Although evidence suggests that blood pressure genetics is similar across ethnic ancestries,^21^ the ethnic diversity of the sample might influence our findings. In addition, sex differences in the genetic architecture exist for related traits.^61^ Thus, sex differences in the genetic architectures of brainstem volumes and blood pressure traits could potentially influence our results. Finally, the cross-sectional design and genetic analyses do not allow causality assessments. Thus, whether brainstem structure influences blood pressure or *vice versa* remains to be clarified. However, it is noteworthy that we found significant associations between brainstem volumes and blood pressure, also when excluding those with hypertension. This suggests that hypertension-related atrophy is not the primary mechanism underlying the significant relationships between brainstem structure and human blood pressure. Longitudinal volumetric studies of individual brainstem nuclei and blood pressure traits are needed to elucidate these relationships further.

In conclusion, we discovered phenotypic and genetic relationships with small effect sizes between the structure of brainstem regions and blood pressure in humans. The overlapping loci implicated genes involved in neuronal development pathways, metal ion transport, and cell-matrix adhesions, thus providing new insights into the mechanisms of the brainstem’s roles in blood pressure regulation and hypertension.

## Data Availability

The UK Biobank resource is open for eligible researchers upon application (http://www.ukbiobank.ac.uk/register-apply/). We made use of publicly available resources for processing the image data and for conducting statistical analyses. We extracted data from individual UK biobank baskets using the *ukb_helper.py* script (https://github.com/precimed/ukb). The project R-scripts for analyzing brainstem volumes and blood pressure traits are available upon publication. We performed the genetic overlap analyses with publicly available methods.^22,24,25^

## Acknowledgments

This research has been conducted using the UK Biobank Resource under Application Number 27412. We performed this work on the *Services for sensitive data* (TSD), University of Oslo, Norway, with resources provided by UNINETT Sigma2 - the National Infrastructure for High-Performance Computing and Data Storage in Norway.

---

**Figure S2:**
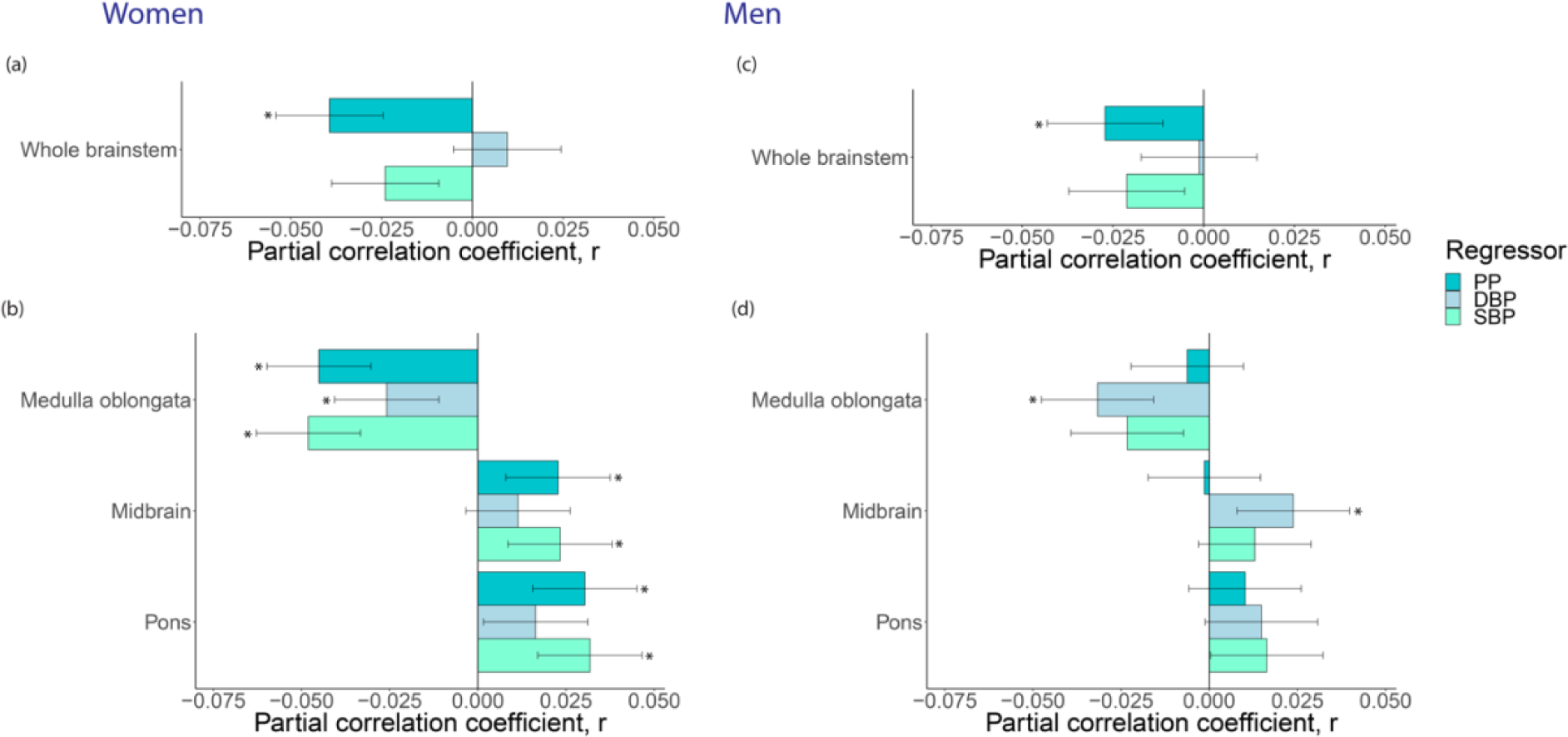
Sex-specific patterns of blood pressure and the brainstem. *Notes:* The association between blood pressure metrics and volumes of **(a)** the whole brainstem and **(b)** brainstem regions in women (n=17,561); and **(c)** the whole brainstem and **(d)** brainstem regions in men (n=15,105). We adjusted for covariates as outlined in the Methods section. *Abbreviations: PP – pulse pressure; DBP – diastolic blood pressure; SBP – systolic blood pressure*.

