## Supplementary information for "Large-scale brainstem neuroimaging and genetic analyses provide new insights into the neuronal mechanisms of hypertension"

### Supplemental Tables

#### Table S1: Demographic and clinical data.

| Variables | Men (n=15,105) | Women (n=17,561) | test | p-value |
|---|---|---|---|---|
| **Age (year)[1]** | 63.6±7.6 | 62.5±7.3 | 12.4 | 2.57215235794614E−35 |
| **Age range (year)** | [44 82] | [45 81] | | |
| **European ancestry, N (%)[2]** | 14619 (96.8) | 17073 (97.2) | 5.2 | 0.0219558911518701 |
| **Systolic blood pressure[1]** | 141±17.1 | 134.8±19 | 30.8 | 6.94772490004767E−205 |
| **Diastolic blood pressure[3]** | 80.6±9.8 | 76.9±9.9 | 33.7 | 2.11479993722548E−244 |
| **Pulse Pressure[1]** | 60.4±13.4 | 57.9±14.8 | 15.7 | 4.27257634612144E−55 |
| **Smoker, N (%)[2]** | 627 (4.2) | 505 (2.9) | 39.1 | 4.03529933395851E−10 |
| **Smoker Current/previous/never** | 627 / 5392 / 9086 | 505 / 5373 / 11683 | | |
| **Alcohol drinker, N (%)[2]** | 14294 (94.6) | 16236 (92.5) | 62.6 | 2.58152078849795E−15 |
| **Alcohol drinker Current/previous/never** | 14294 / 471 / 340 | 16236 / 607 / 718 | | |
| **Height (cm)[1]** | 176.2±6.6 | 162.8±6.2 | 186.8 | 0 |
| **Weight (kg)[1]** | 83.4±13.2 | 68.9±12.9 | 99.9 | 0 |
| **Body mass index[1]** | 26.8±3.8 | 26±4.7 | 17.8 | 8.24164917491708E−71 |
| **Waist circumference (cm)[1]** | 93.5±10.4 | 82.3±11.5 | 93 | 0 |
| **Hip circumference (cm)[1]** | 100.5±7.2 | 100.7±9.7 | −2 | 0.0484740227019555 |
| **Waist-to-hip ratio[1]** | 0.9±0.1 | 0.8±0.1 | 157.1 | 0 |
| **Diabetic[2]** | 400 (2.6) | 226 (1.3) | 79.3 | 5.27538194434006E−19 |
| **High cholesterol[2]** | 2556 (16.9) | 1773 (10.1) | 328.5 | 2.07813962016952E−73 |
| **Hypertension[2]** | 3974 (26.3) | 3197 (18.2) | 310.8 | 1.46474133078869E−69 |

Notes: Report mean ± standard deviation for continuous variables. P-values≤0.05 are considered significant. A p-value of zero indicates that the p-value is below numeric precision.

[1] Welch Two Sample t-test
[2] Pearson's Chi-squared test with Yates' continuity correction
[3] Two Sample t-test

### Table S2: T-tests of the whole brainstem and brainstem regions split on sex.

| Variables | Men (n=15,105) | Women (n=17,561) | test | p-value |
|---|---|---|---|---|
| **Medulla oblongata (ml)** [1] | 4.2±0.6 | 3.8±0.5 | 62.6 | 0 |
| **Pons (ml)** [1] | 15.2±1.9 | 13.7±1.7 | 73.2 | 0 |
| **Midbrain (ml)** [1] | 6.4±0.6 | 5.7±0.5 | 108.5 | 0 |
| **Whole brainstem (ml)** [1] | 26.1±3 | 23.5±2.6 | 82.6 | 0 |
| **Intracranial volume (ml)** [1] | 1579.9±135.2 | 1413.9±115 | 118.4 | 0 |

*Notes:* A p-value of zero indicates that the p-value is below numeric precision.

[1] Welch Two Sample t-test.

### Supplemental Figures

#### Figure S1: Density plots demographic variables.

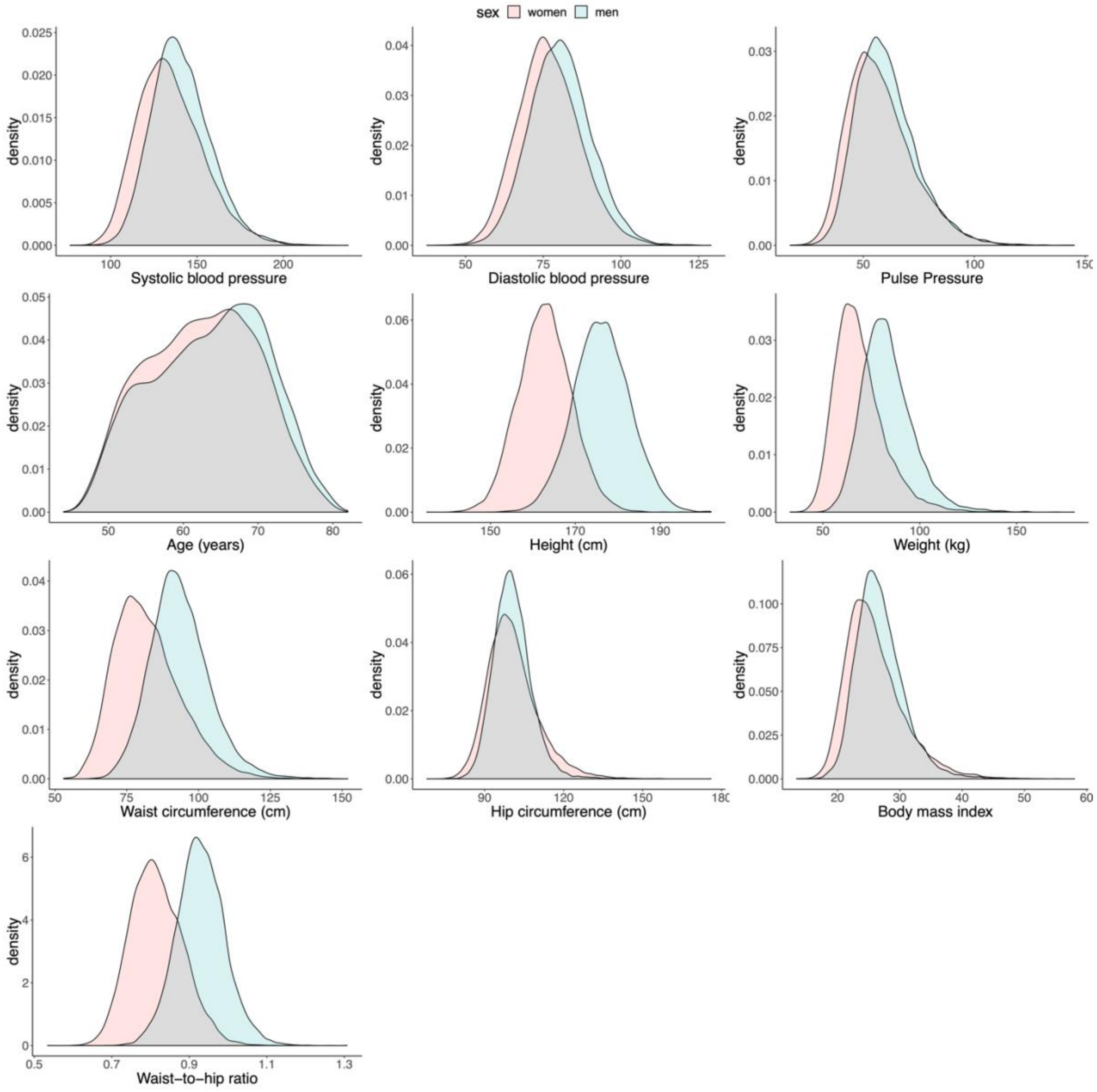

#### Figure S2: Density plots of brain magnetic resonance imaging (MRI) variables.

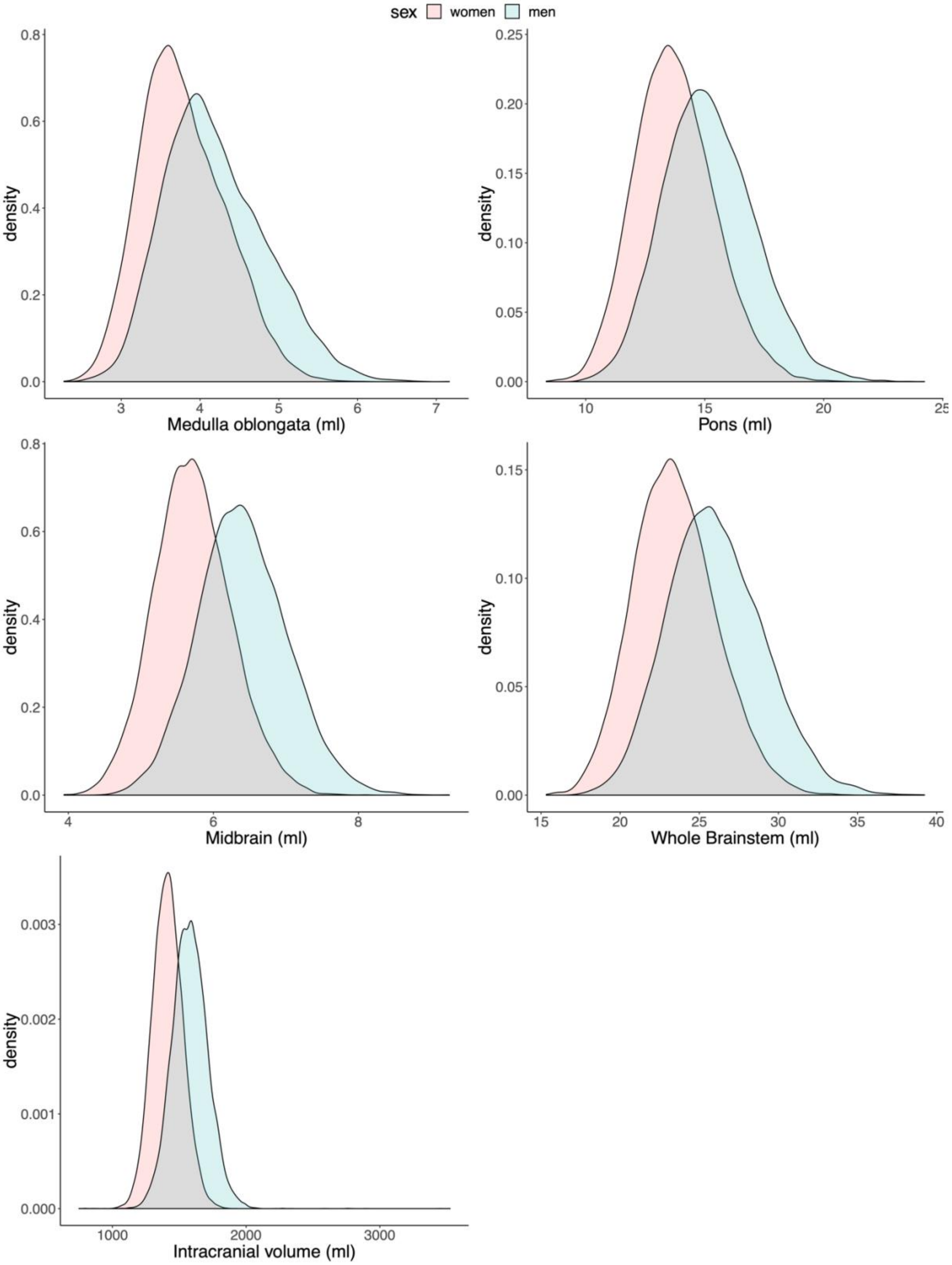

*Notes:* All brain structures are given in ml.

**Figure S3: Conditional Q-Q and log-likelihood plots for systolic blood pressure and brainstem regions.**

(a) Systolic blood pressure & the whole brainstem

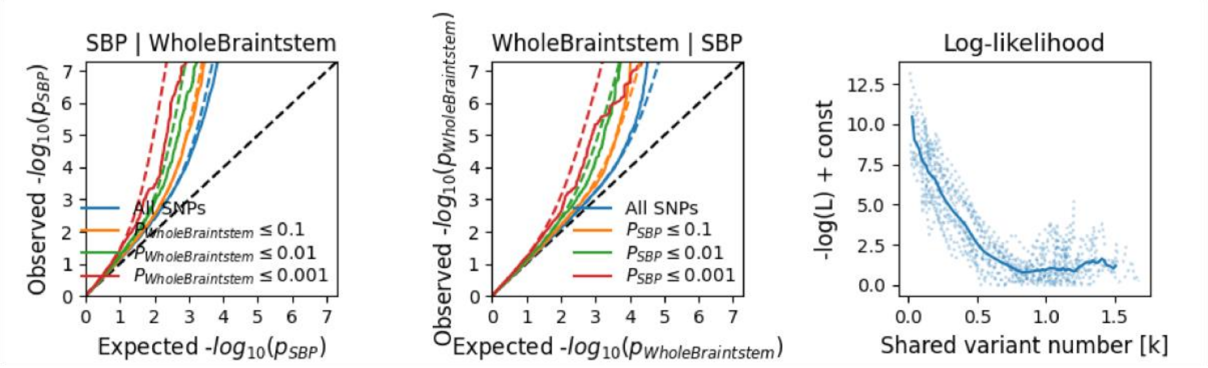

(b) Systolic blood pressure & Medulla Oblongata

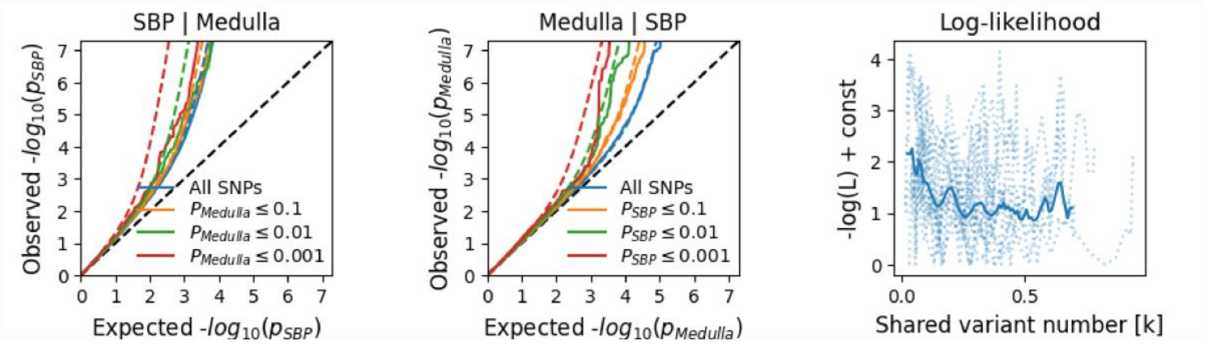

(c) Systolic blood pressure & Midbrain

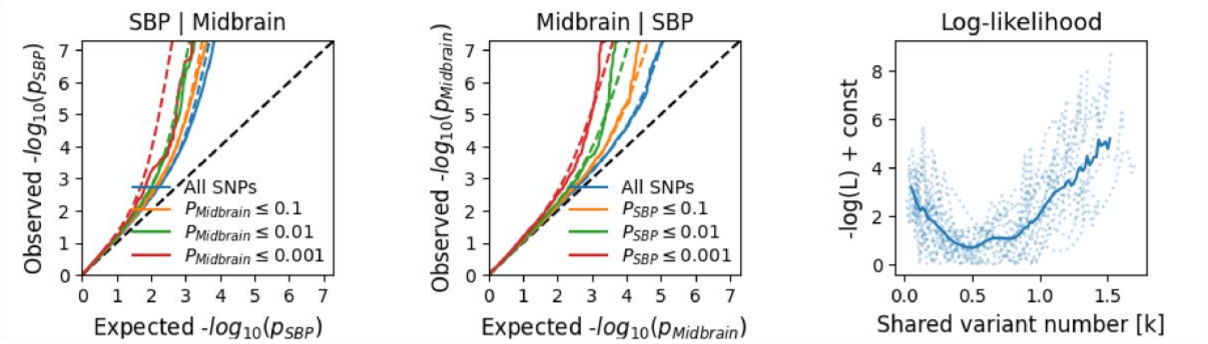

(d) Systolic blood pressure & Pons

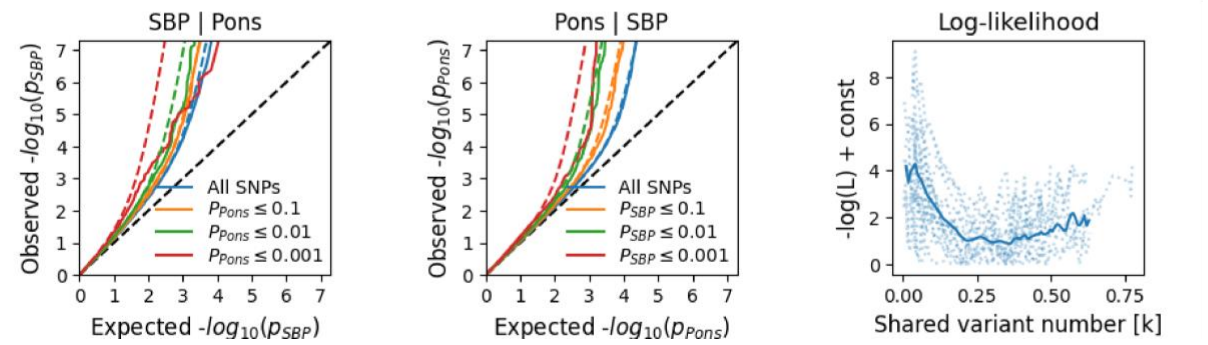

*Abbreviations:* Medulla – Medulla oblongata; SBP – systolic blood pressure; Q-Q – quantile-quantile.

**Figure S4: Conditional Q-Q and log-likelihood plots for diastolic blood pressure and brainstem regions.**

(a) Diastolic blood pressure & the whole brainstem

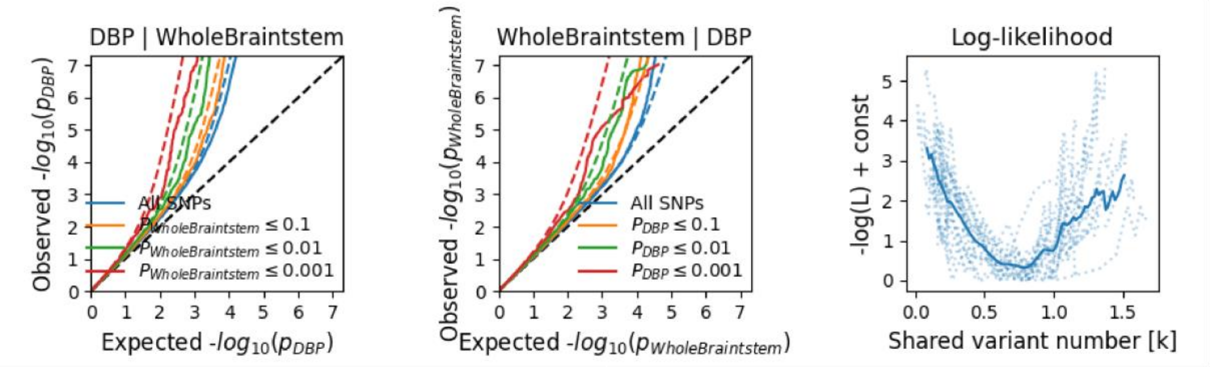

(b) Diastolic blood pressure & Medulla Oblongata

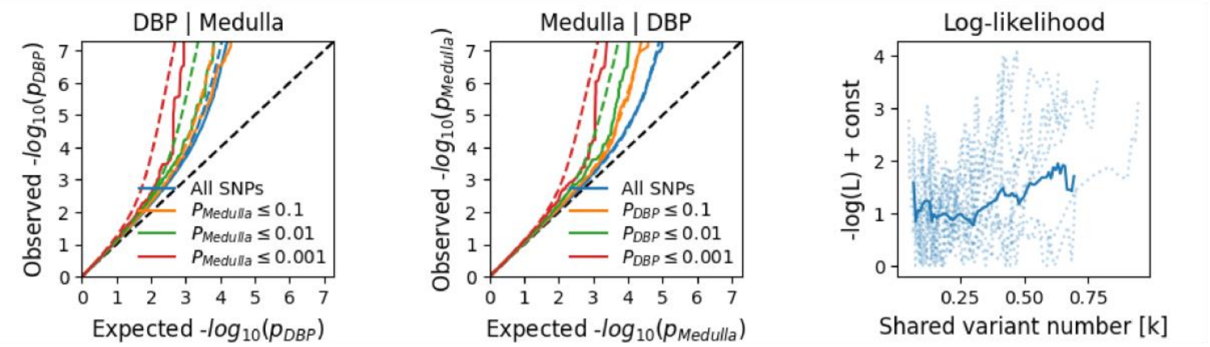

(c) Diastolic blood pressure & Midbrain

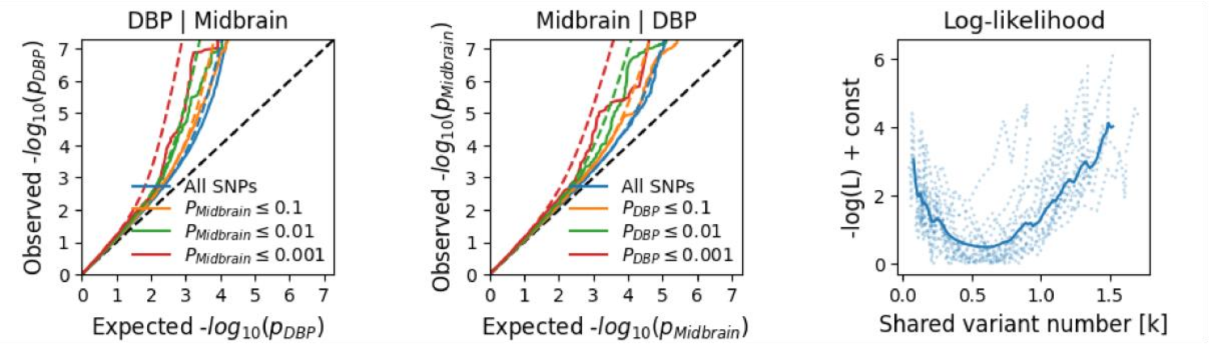

(d) Diastolic blood pressure & Pons

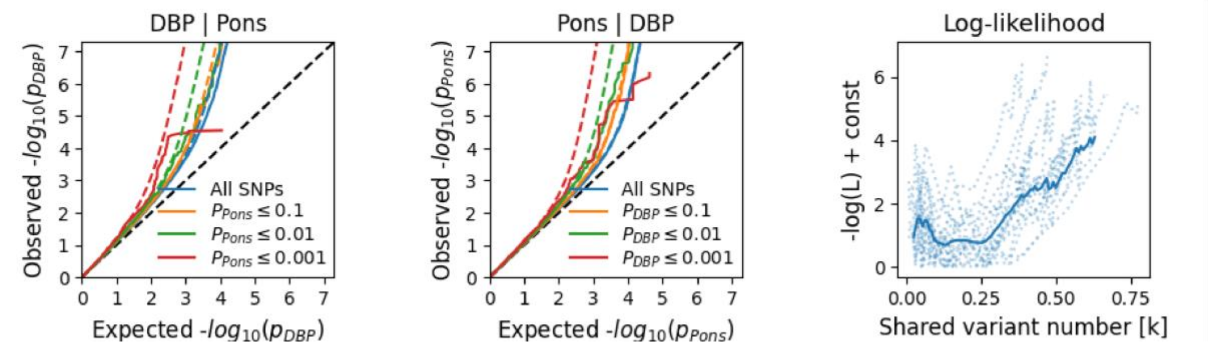

*Abbreviations:* DBP – diastolic blood pressure; Medulla – Medulla oblongata; Q-Q – quantile-quantile.

**Figure S5: Conditional Q-Q and log-likelihood plots for pulse pressure and brainstem regions.**

(a) Pulse pressure & the whole brainstem

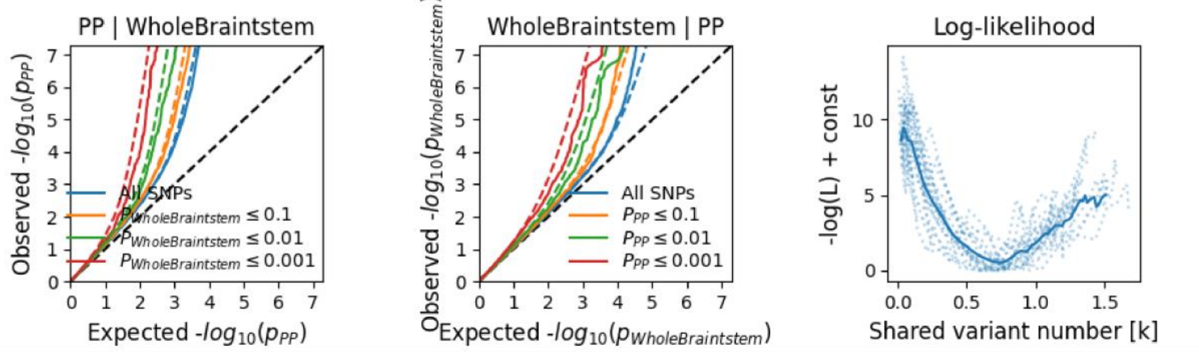

(b) Pulse pressure & Medulla Oblongata

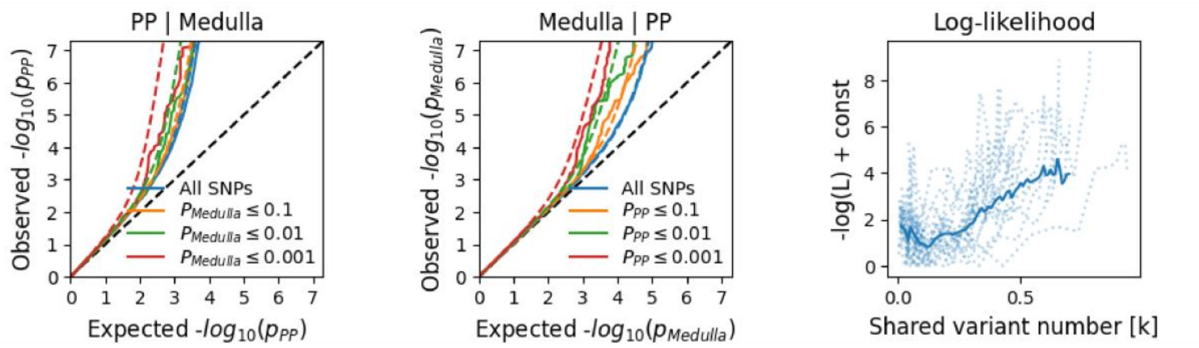

(c) Pulse pressure & Midbrain

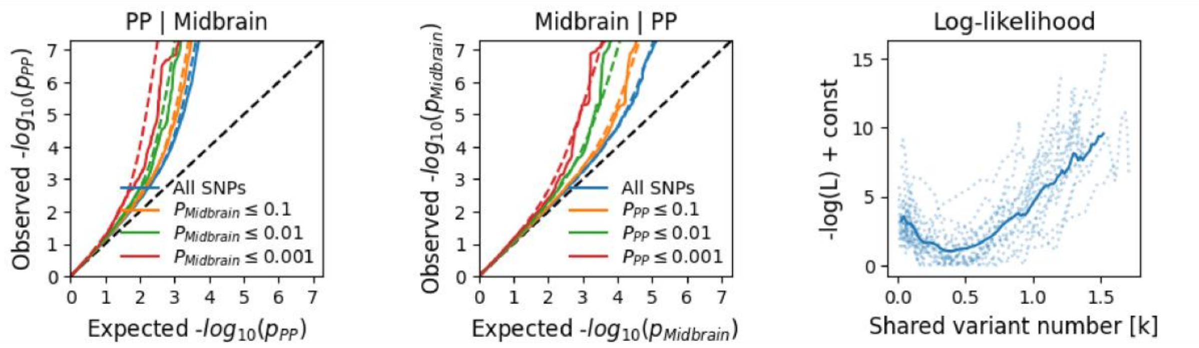

(d) Pulse pressure & Pons

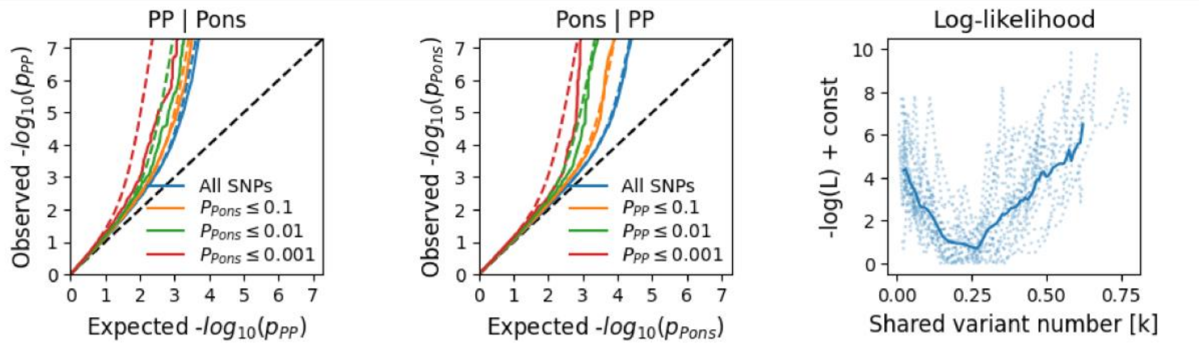

*Abbreviations:* Medulla – Medulla oblongata; PP – pulse pressure; Q-Q – quantile-quantile.

**Figure S6: Blood pressure and brainstem volumes in a non-hypertensive state.**

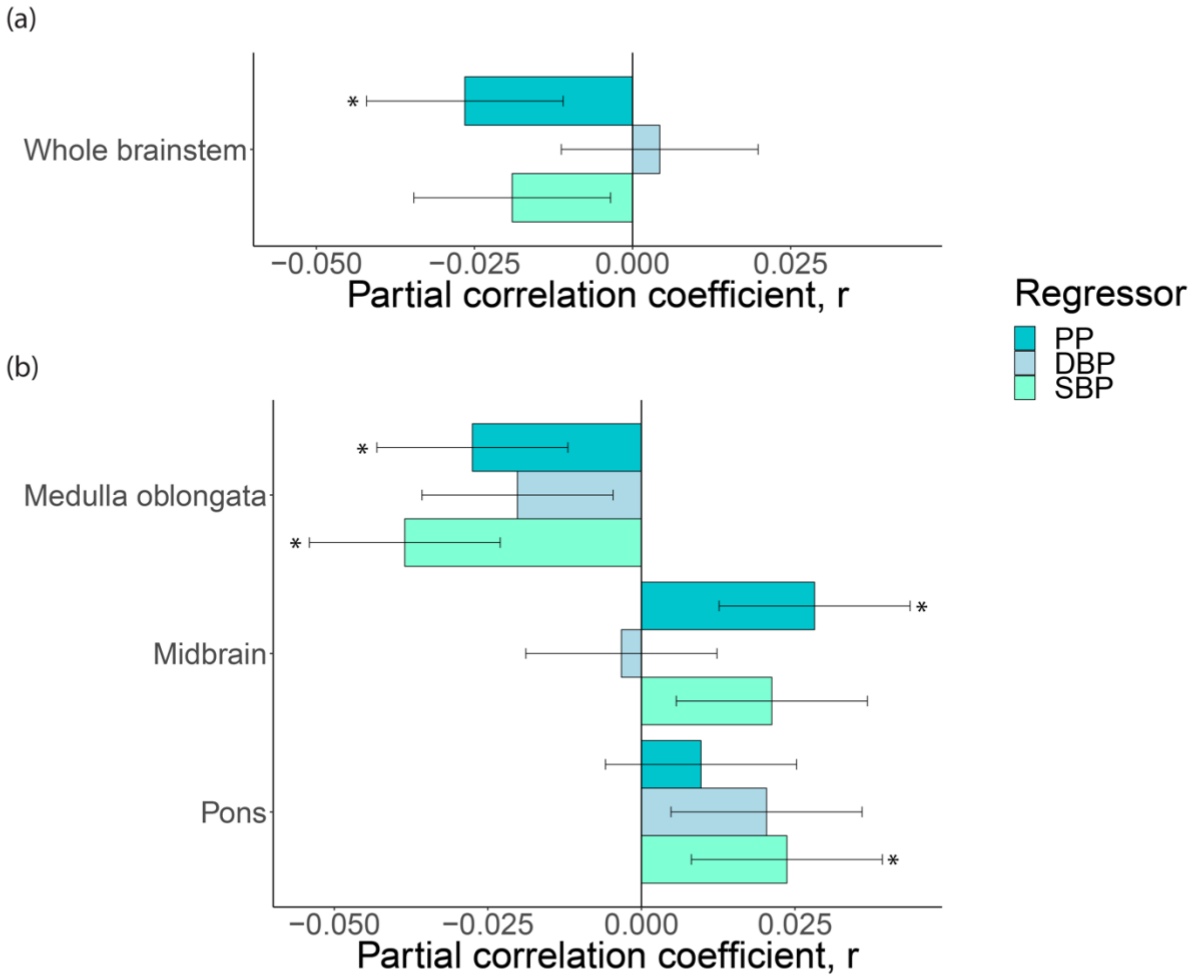

*Notes:* In participants without hypertension (n=15,870), the association between blood pressure metrics and volumes of the **(a)** whole brainstem and **(b)** brainstem regions. We adjusted for covariates as outlined in the Methods section. *Abbreviations: DBP – diastolic blood pressure; PP – pulse pressure; SBP – systolic blood pressure.*

### Figure S7: Age-specific patterns of blood pressure and the brainstem.

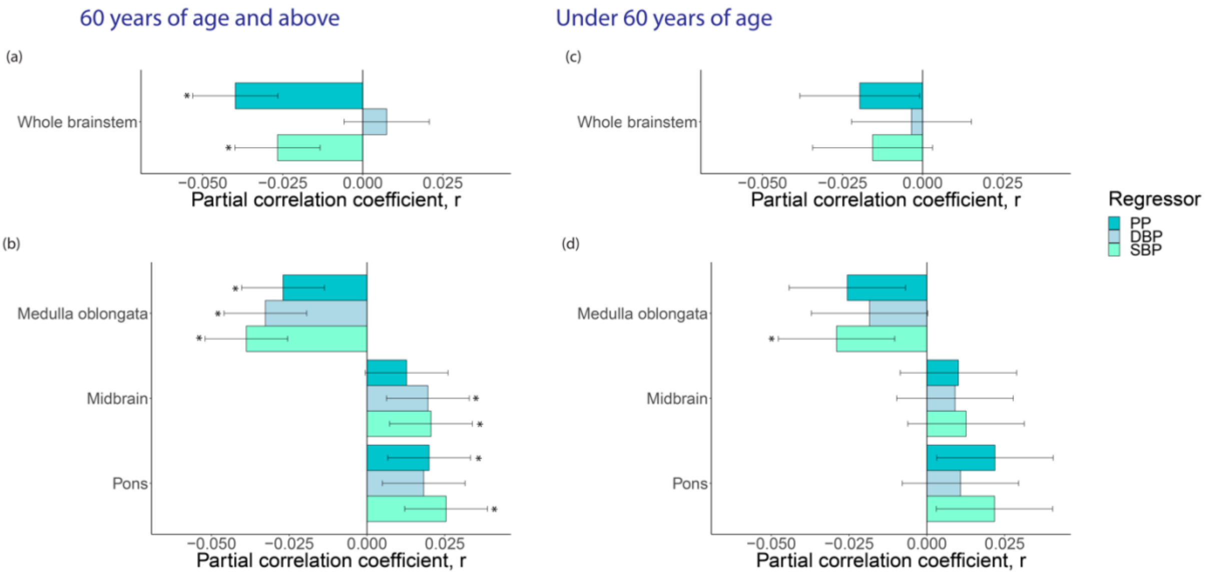

*Notes:* The association between blood pressure metrics and volumes of **(a)** the whole brainstem and **(b)** brainstem regions in those ≥60 years of age (n=21,697); and **(c)** the whole brainstem and **(d)** brainstem regions in those <60 years of age (n=10,969). We adjusted for covariates as outlined in the Methods section. *Abbreviations: DBP – diastolic blood pressure; PP – pulse pressure; SBP – systolic blood pressure.*

**Figure S8: The genetic correlation between traits from LDScore regression.**

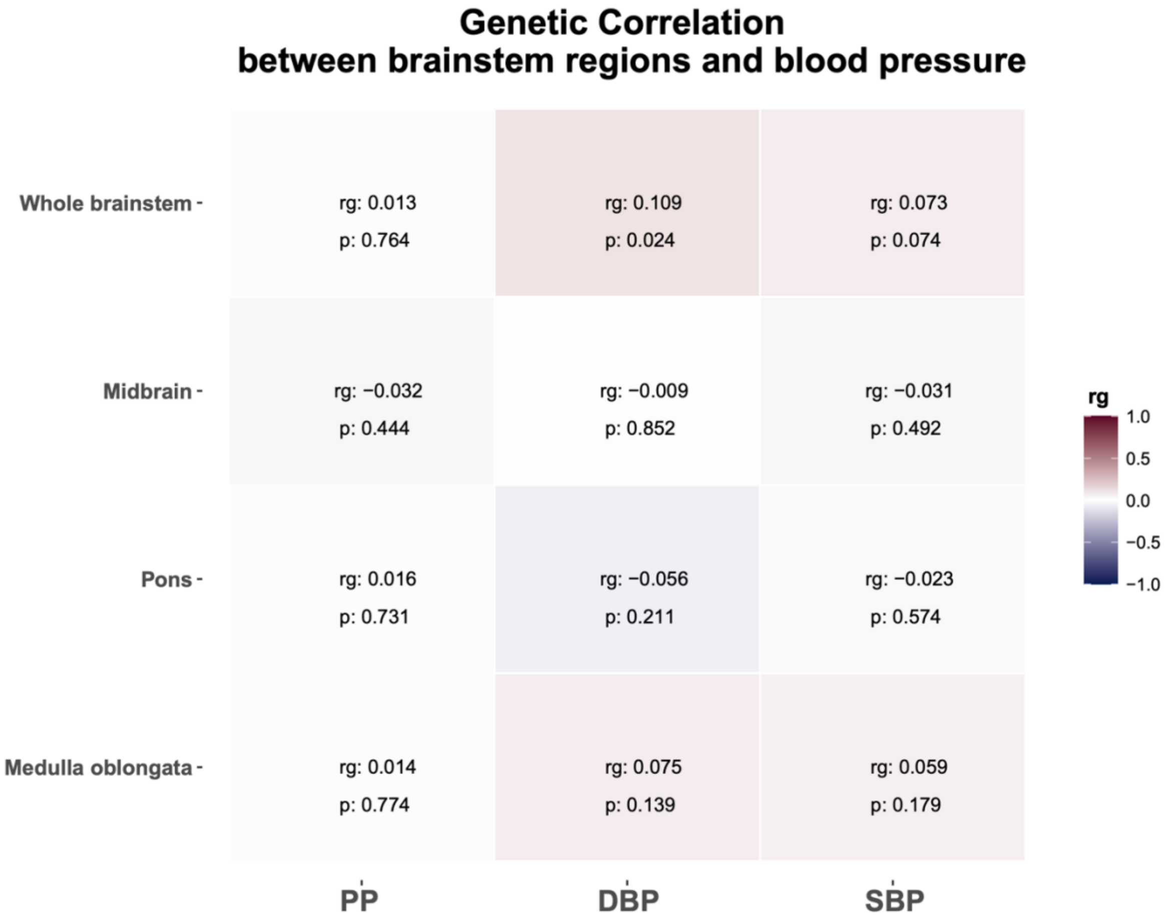

*Abbreviations: PP – pulse pressure; DBP – diastolic blood pressure; SBP – systolic blood pressure; Whole – whole brainstem.*

**Figure S9: Q-Q plots of the genetic overlap analyses: systolic blood pressure & brainstem volumes.**

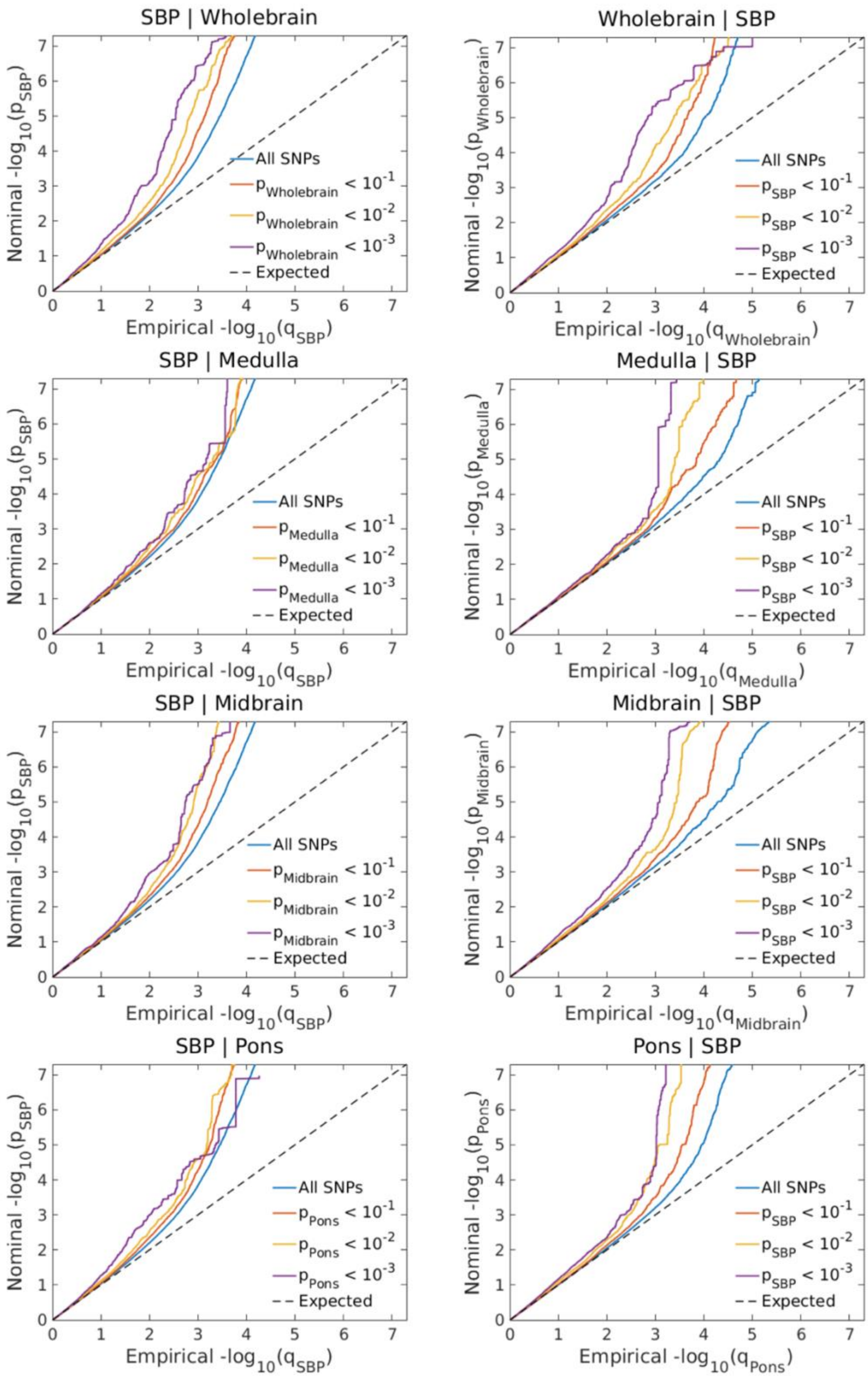

*Abbreviations: Medulla – Medulla Oblongata; SBP – systolic blood pressure; Wholebrain – whole brainstem; Q-Q – quantile-quantile.*

**Figure S10: Q-Q plots of the genetic overlap analyses: diastolic blood pressure & brainstem volumes.**

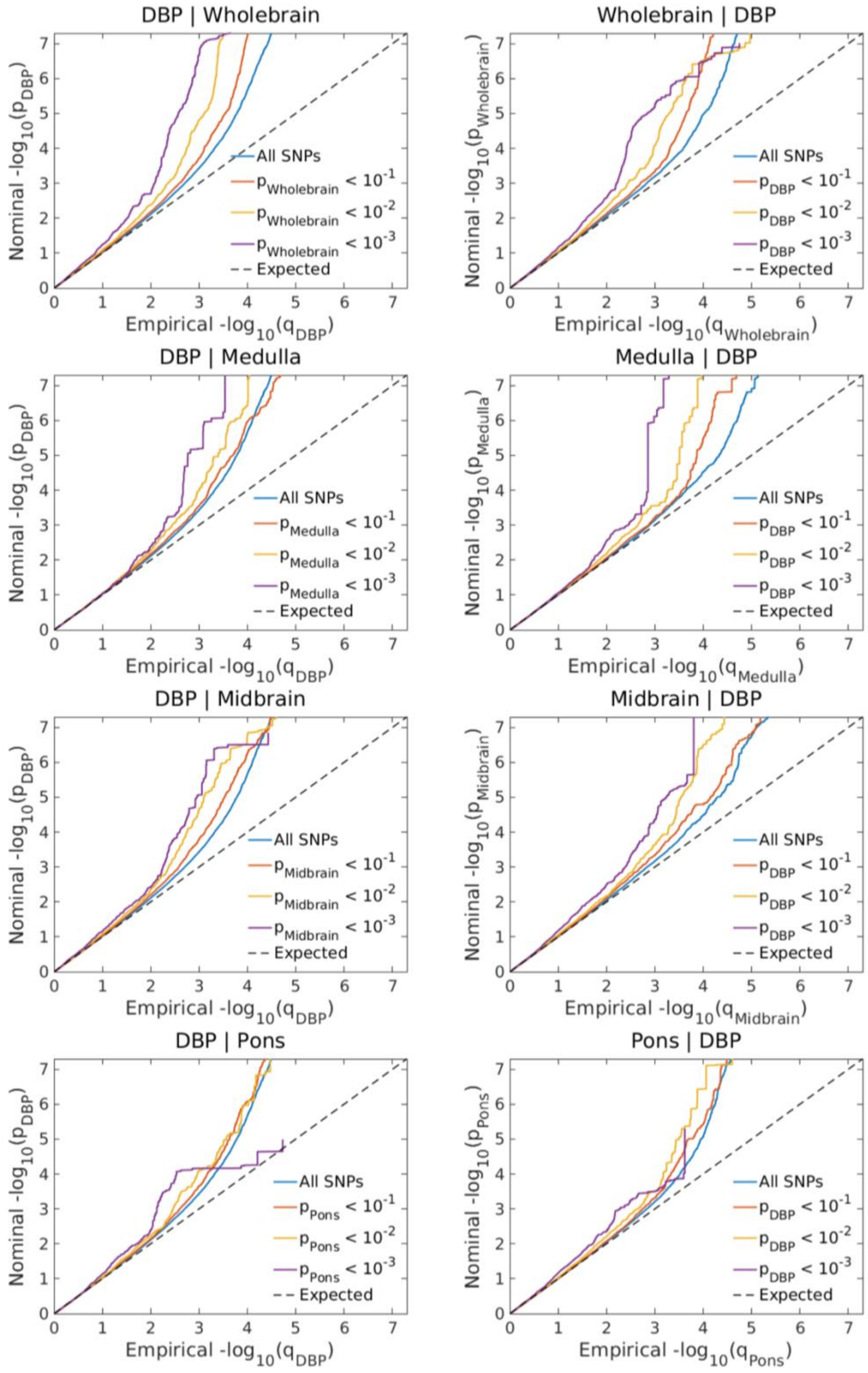

*Abbreviations: Medulla – Medulla Oblongata; SBP – systolic blood pressure; Wholebrain – whole brainstem; Q-Q – quantile-quantile.*

#### Figure S11: Q-Q plots of the genetic overlap analyses: pulse pressure & brainstem volumes.

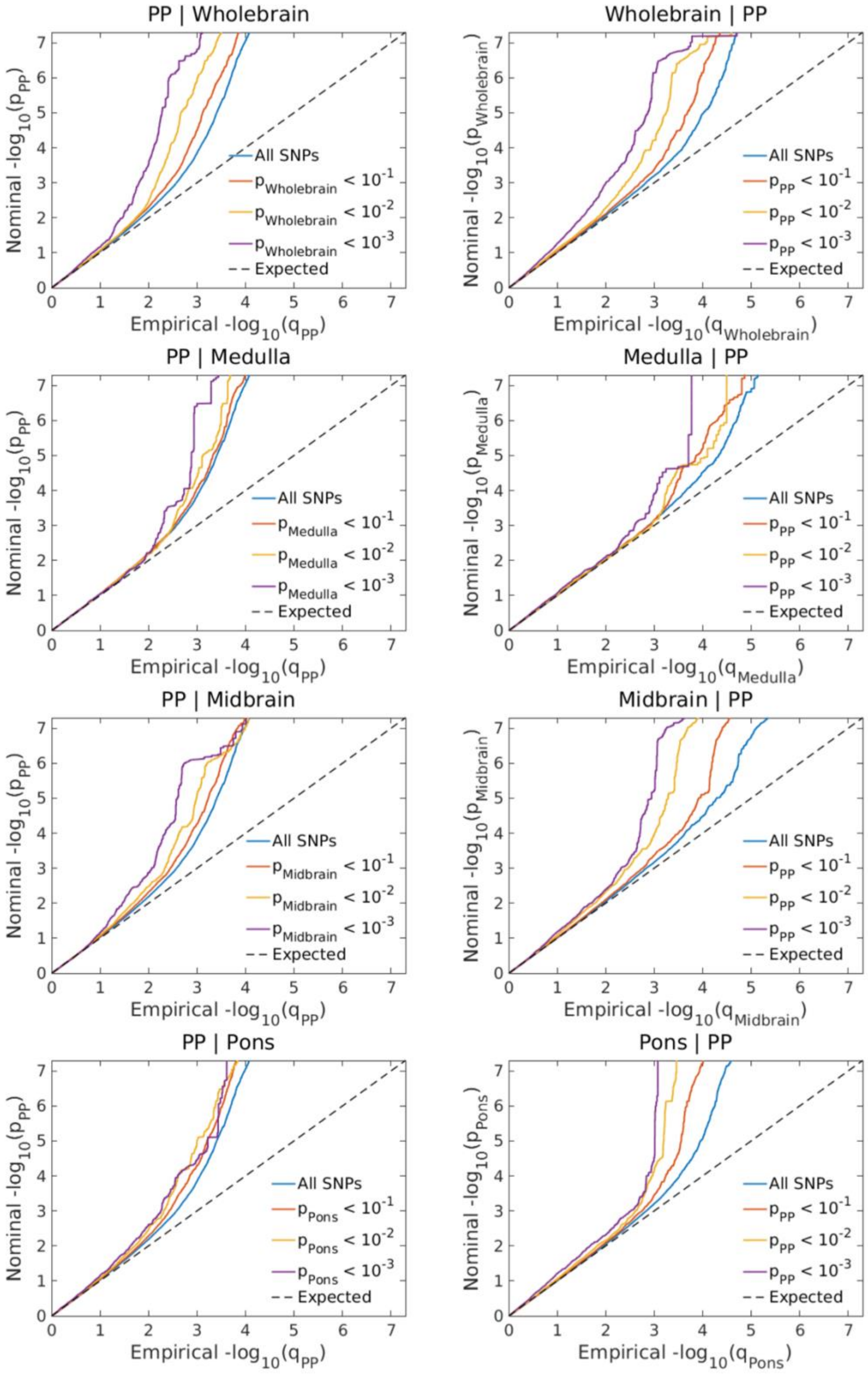

*Abbreviations: Medulla – Medulla Oblongata; PP - pulse pressure; Wholebrain – whole brainstem; Q-Q – quantile-quantile.*

### Supplemental Notes

#### Note S1: Overview of extracted demographic and clinical data.

Table SN1 yields an overview of the extracted demographic and clinical data from the UK Biobank.

We created binary (yes/no) variables for self-reported diagnosis (data-field 20002.2.*) of diabetes (diabetes, diabetes type 1, diabetes type2), hypertension, and high cholesterol (hypercholesterolemia); and binary variables for current alcohol consumption and cigarette smoking (yes: current, no: previous/never). When available, we created a binary variable for self-identified European/non-European ancestry based on the MRI time point. We complemented missing ancestry data at the MRI time point with baseline information since it does not change with time (although knowledge or perception of ethnic ancestry can change).

**Table SN1: Extracted demographic and clinical variables with data-field ID.**

| Data-field ID | Field Description |
|---|---|
| 31.0.0 | Sex |
| 54.2.0 | Assessment center |
| 21003.2.0 | Age |
| 50.2.0 | Standing height |
| 21002.2.0 | Weight |
| 48.2.0 | Waist circumference |
| 49.2.0 | Hip circumference |
| 21001.2.0 | BMI |
| 21000.*.0[1] | Ethnicity |
| 20002.2.*[2] | Non-cancer diagnosis (self-reported) |
| 20117.2.0 | Alcohol drinking status |
| 20116.2.0 | Cigarette smoking status |
| 4080.2.*[3] | Systolic blood pressure – automatic reading |
| 93.2.*[3] | Systolic blood pressure – manual reading |
| 4079.2.*[3] | Diastolic blood pressure – automatic reading |
| 94.2.*[3] | Diastolic blood pressure – manual reading |

[1] Baseline and imaging timepoint extracted.

[2] All sub-items (indicated by *), from 0 to 32 extracted.

[3] Computed the average from two subsequent readings (indicated by *), if available. Otherwise, used single reading.

### Note S2: Brainstem regions without adjusting for the whole brainstem.

In this supplemental note, we present the results concerning volumes of brainstem regions without including adjustment for the whole brainstem in the regression model. We adjusted for all other covariates as presented in the Methods section.

#### Brainstem regions and blood pressure metrics

We found significant negative associations between medulla oblongata volumes and systolic blood pressure (r=-0.04, p=1.4×$10^{-12}$) and pulse pressure (r=-0.04, p=1.4×$10^{-13}$). Pons volume was significantly associated with pulse pressure (r=-0.03: p=1.0×$10^{-6}$) and systolic blood pressure (r=-0.02, p=0.0035), and similarly, midbrain volume was associated with pulse pressure (r=-0.02, p=2.6×$10^{-5}$) (**Figure SN1, Table S3a**).

***Figure SN1: Blood pressure and brainstem regions.***

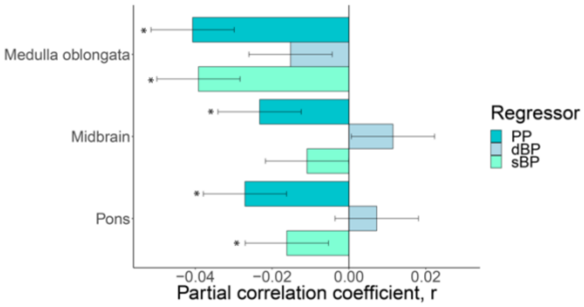

*Notes:* The association between blood pressure metrics and brainstem regions (n=32,666) without adjustment for the whole brainstem. *Abbreviations: dBP – diastolic blood pressure; PP – pulse pressure; sBP – systolic blood pressure.*

### Brainstem regions and blood pressure metrics for non-hypertensive state

In follow-up analyses of participants without self-reported or derived hypertension (n=15,870; 61.4% women), we observed highly similar association patterns as in the full sample. Significant associations were limited to the medulla oblongata and systolic blood pressure (r=-0.04, $p=8.9\times10^{-7}$) and pulse pressure (r=-0.04, $p=1.8\times10^{-6}$), and pons and pulse pressure (r=-0.02, p=0.0030) (**Figure SN2; Table S4a**).

***Figure SN2: Blood pressure and brainstem regions for the non-hypertensive state.***

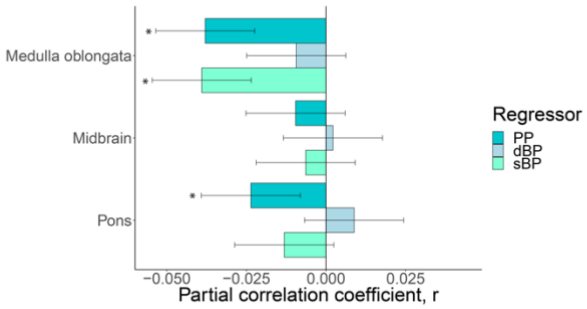

*Notes:* The association between blood pressure metrics and brainstem regions (n=15,870) in participants without hypertension without adjustment for the whole brainstem. *Abbreviations: dBP – diastolic blood pressure; PP – pulse pressure; sBP – systolic blood pressure.*

### Age-related patterns

For those 60 and above (n=21,697; 52.2% women), there were significant findings for medulla oblongata and systolic blood pressure (r=-0.05, p=3.3×$10^{-11}$) and pulse pressure (r=-0.05, p=1.7×$10^{-12}$). For pons volume, we observed a significant association with pulse pressure (r=-0.03, p=4.7×$10^{-7}$) and systolic blood pressure (r=-0.02, p=0.0030) and similarly between midbrain volume and pulse pressure (r=-0.03, p=1.5×$10^{-5}$). There were no significant findings for diastolic blood pressure (**Figure SN3a; Table S5a**)

There were fewer significant associations for those under 60 (n=10,969; 57.0% women) compared to the older age group but a similar pattern. We only observe a significant association between medulla oblongata volume and pulse pressure (r=-0.03, p=0.0010) and systolic blood pressure (r=-0.03: p=0.0015) (**Figure SN3b; Table S**6a).

***Figure SN3: Age-specific patterns of blood pressure and brainstem regions.***

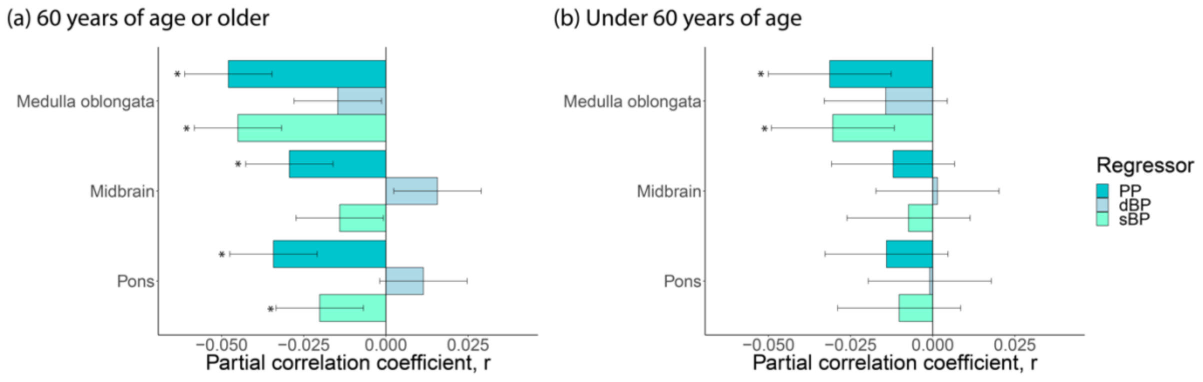

*Notes:* The association between blood pressure metrics and brainstem regions in **(a)** those ≥60 years of age (n=21,697) and **(b)** those <60 years of age (n=10,969) without adjusting for the whole brainstem. *Abbreviations: dBP – diastolic blood pressure; PP – pulse pressure; sBP – systolic blood pressure.*

### Sex-related patterns

For women (n=17,561), we observed significant associations between the medulla oblongata and systolic blood pressure (r=-0.05, p=7.7×$10^{-11}$) and pulse pressure (r=-0.06, p=5.1×$10^{-15}$). Midbrain and pons volumes was significantly associated with pulse pressure (Midbrain: r=-0.02, p=0.0018; Pons: r=-0.03; p=3.2×$10^{-5}$) (**Figure SN4a, Table S7a**).

For men (n=15,105), the significant associations were limited to medulla oblongata volume and systolic blood pressure (r=-0.03, p=0.0001) and pulse pressure (r=-0.03, p=0.0020). Pons and Midbrain volumes were both significantly associated with pulse pressure (Pons: r=-0.02, p=0.0030; Midbrain r=-0.02, p=0.0024) (**Figure SN4b, Table S8a**).

***Figure SN4: Sex-specific patterns between blood pressure and brainstem regions.***

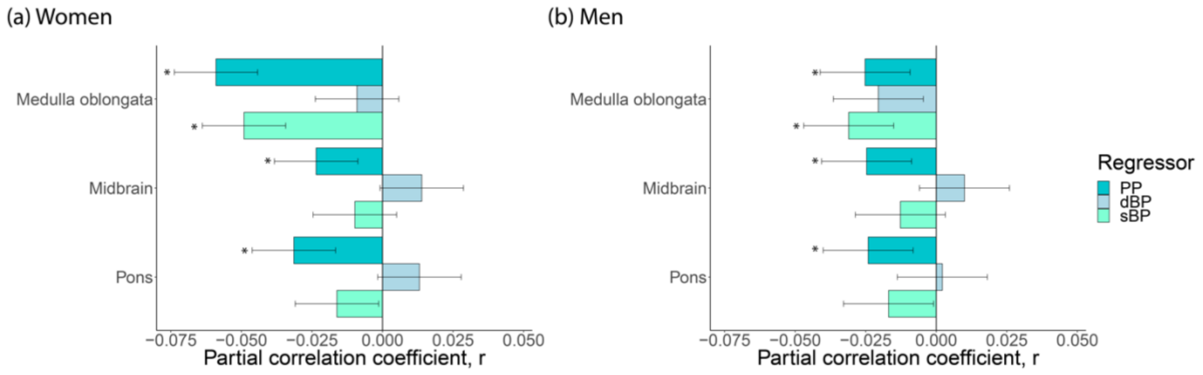

*Notes:* The association between blood pressure metrics and brainstem regions in **(a)** women (n=17,561) and **(b)** men (n=15,105) without adjusting for the whole brainstem. *Abbreviations: dBP – diastolic blood pressure; PP – pulse pressure; sBP – systolic blood pressure.*
